# Neural Correlates of Eye Contact and Social Function in Autism Spectrum Disorder

**DOI:** 10.1101/2021.10.01.21264368

**Authors:** Joy Hirsch, Xian Zhang, J. Adam Noah, Swethasri Dravida, Adam Naples, Mark Tiede, Julie M. Wolf, James C. McPartland

**Affiliations:** Brain Function Laboratory, Department of Psychiatry, Yale School of Medicine, 300 George St., Suite 902, New Haven, CT, USA; Interdepartmental Neuroscience Program, Yale School of Medicine, New Haven, CT, 06511, USA; Yale Child Study Center, Nieson Irving Harris Building, 230 South Frontage Road, Floor G, Suite 100A, New Haven, CT, 06519, USA; Department of Neuroscience, Yale School of Medicine, New Haven, CT, 06511, USA; Department of Comparative Medicine, Yale School of Medicine, New Haven, CT, 06511, USA; Department of Medical Physics and Biomedical Engineering, University College London, London, WC1E 6BT, UK; Haskins Laboratories, 300 George St., Suite 900, New Haven, CT, 06511, USA

**Keywords:** autism spectrum disorder (ASD), live eye-to-eye contact, interactive face processing, dorsal stream, functional near-infrared spectroscopy (fNIRS), hyperscanning, social symptomatology, visual sensing

## Abstract

Reluctance to make eye contact during natural interactions is a central diagnostic criterion for autism spectrum disorder (ASD). However, the underlying neural correlates for natural eye contacts in ASD are unknown, and diagnostic biomarkers are active areas of investigation. Here, neuroimaging, eye-tracking, and pupillometry data were acquired simultaneously using two-person functional near-infrared spectroscopy (fNIRS) during live eye-to-eye contact and eye-gaze at a video face in typically developed (TD) and ASD participants to identify the neural correlates of live eye-to-eye contact in both groups. Direct comparisons between ASD and TD participants showed decreased right dorsal parietal activity and increased right ventral temporal-parietal activity for ASD relative to TD during live eye-to-eye contact (p≤0.05, FDR-corrected) consistent with the hypothesis of alternative neural systems for live eye contact. The additional hypothesis that hypoactivity of the right dorsal-parietal regions during eye contact is associated with social performance in ASD was supported by the correlation of right dorsal parietal activity with individual measures of social function: ADOS-2, Autism Diagnostic Observation Schedule, 2^nd^ Edition (r = -0.69); and SRS-2, Social Responsiveness Scale, Second Edition (r = -0.58). That is, as social ability decreased, the neural responses in the right dorsal parietal region to real eye-contact also decreased consistent with a neural correlate for social characteristics in ASD.

## Introduction

Eye contact with another human is an impactful and fundamental component of in-person social behavior. Intermittent eye-to-eye fixations are widely thought of as particularly potent stimuli. For example, proverbial claims that “the eyes are the window to the soul” are taken as self-evident, and eye-to-eye contact is attributed with literary properties such as the “spark” that ignites a “social connection” and the sharing of information. Wisdom regarding the social significance of eye gaze is long-standing in classical literature and may have its source in a quote from Marcus Tullius Cicero‘s *Orator*: “*Ut imago est animi voltus sic indices oculi”* translated as, “For as the face is the image of the soul, so are the eyes its interpreters” (1). Although there are cultural variations related to the interpretation of interpersonal eye gaze (2–5), eye contacts universally signal social cues such as levels of engagement, emotional status, intention, judgment, and an array of nuanced exchanges of social information including an invitation for interaction. Direct eye contact with another person is conventionally taken as a significant interaction, regardless of the cultural norms. Given the salience of real face-to-face and dynamic eye-to-eye interactions, the development of two-person methodologies and a theoretical framework for understanding the neural biology of live face-to-face related social cues is a high priority.

Autism spectrum disorder (ASD) is a complex neurodevelopmental condition including distinct behavioral, communicative, and social responses such as reluctance to make eye contact during natural social interactions (6). The underlying neurobiology of ASD is typically investigated by single non-interactive participants and, therefore, the biological underpinnings of observed behavioral differences during live and interactive social processes are not well understood. The single participant focus of conventional neuroimaging is a contributing factor to this knowledge gap, as the neural systems that underlie social differences in ASD cannot be directly investigated in passive stimulus-response paradigms. Nonetheless, single participant stimulation paradigms have contributed the foundation for the basic neuroscience of social ability in ASD.

For example, in the case of visual sensing and low-level visual processes, variations in patterns of eye-gaze are well documented in ASD (7–15). Reduced responses to emotional cues conveyed by simulated facial dynamics (16) and reduced production of facial expressions that signal emotional content (17–20) have also been reported. Reduced occipital pole responses to pictured eyes for ASD participants observed by fMRI suggest atypical early visual processing (21). Eye gaze at pictured eyes with emotional content has been associated with abnormally high activation in subcortical systems including superior colliculus, pulvinar nucleus of the thalamus, and the amygdala. Elevated amygdala responses to neutral faces have also been associated with eye gaze suggestive of increased arousal for face stimuli (22).

Electroencephalography (EEG) findings in ASD show increased latency of event-related potentials (ERPs) to pictured eye stimuli relative to typically developed (TD) participants (23, 24). This latency has been shown to be related to gaze directed at the eyes of a pictured face in TD individuals (Parker et al., 2021). Atypical and reduced responses to simulated faces and robots have also been shown using fNIRS in support of the hypothesis of alternative neural pathways for face processing (25).

Temporal cortex is activated specifically during viewing of eye movements in the TD population as shown by fMRI and electrophysiology and confirmed by primate neurophysiology (26–29). In contrast, in ASD, hypoactivation of superior temporal sulcus and face-related dorsal areas consisting of the somatosensory and premotor cortex previously associated with face processing have also been reported (30). Atypical brain activation patterns during face-to-face joint attention in adults with ASD revealed reduced signal differentiation between joint attention and control conditions within the superior temporal sulcus and dorsal medial prefrontal cortex (31). Hypoactive high-level cognitive and language systems have also been reported (32, 33). These prior findings of atypical early and higher-order perceptual, cognitive, and language processing are consistent with the overarching hypothesis of impacted social-communicative systems related to face and eye- processing in ASD (34).

In natural real-life situations, humans typically perceive complex facial information conveyed by facial expressions within very brief exposures including subliminal presentations of fearful faces (35). Dynamic and reciprocal face interactions are primary sources of social information and streaming of cues extracted from faces guide real live perceptions and behaviors. However, self-declared autistic adolescents and adults have reported struggling with flexibly and strategically extracting information from the face using eye gaze during face-to-face interactions (36). The importance of investigating real-life facial interactions in ASD has been recognized by recent calls for “second-person neuroscience” (37–39). A theoretical context for this notion in clinical applications has been proposed as “second person neuropsychiatry” (Schilbach, 2016) aimed at the development of quantitative assessments of dyadic social interactions referred to “interaction-based phenotyping” (Schilbach, 2019). However, the paucity of neuroimaging techniques to acquire dyadic information on dynamic face processing during real social interactions has challenged advances in the development of these methods and in understanding the underlying neurobiology of these processes (40, 41) and their variations in ASD.

In response to this knowledge gap, recent developments in functional near-infrared spectroscopy (fNIRS) have applied hyperscanning (simultaneous brain scanning of two individuals during live interactions) to pave the way for much-needed studies of live interactions between individuals and investigations of neural substrates associated with atypical social interactions. Technical advances in fNIRS (42, 43) and the immediate need to understand the biological components of live and interactive human social behaviors have supported the emergence of neuroimaging technology to investigate dynamic face-to-face and eye contact behaviors in ASD.

The current investigation aims to identify neural systems responsive to *in vivo* eye contact in both ASD and TD and to examine their relationship to social performance. The overarching hypothesis is that interactive face processing, with eye contact being a central component, engages complex neural encoding of high-level demands for rapid interpretation of subtle eye movements and facial cues that convey social meaning. These cues are not included in non-interactive and conventional simulated face stimuli. Thus, it is expected that neural processing of interactive faces in ASD and TD groups will include social systems (44); interactive face processing systems (45–47); and motion-sensitive systems (48) hypothesized to be differentiated between TD and ASD groups. Here we test the specific hypothesis that individual differences in social function in ASD are predicted by neural responses associated with live eye-to-eye contact.

TD and ASD adults were compared during real gaze at the eyes of a same-sex lab partner and gaze at a comparable dynamic face video (instructions were to gaze at the eyes under both conditions). In the real-person interaction condition (Real Eye), partners viewed each other’s faces directly while sitting across a table from one other. Findings were compared with a condition in which participants and their lab partners both viewed the eyes of a size-matched face displayed on a video monitor (Video Eye). The contrast between these two conditions and direct comparisons between ASD and TD groups tested the hypothesis that neural processes responsive to real eye-to-eye contact are altered in ASD relative to TD. Eye tracking and pupillometry during these conditions were used to test the related hypothesis that gaze characteristics (visual sensing) and an autonomic indicator of arousal (pupil size variation) also varied during face processing between TD and ASD. The social performance for all ASD participants was assessed by clinical interview, including administration of the Autism Diagnostic Observation Schedule (2^nd^ Edition, ADOS-2) (49). Self-report measures were given to both ASD and TD participants, including the Social Responsiveness Scale (Second Edition, SRS-2) (50). A goal of this investigation was to determine how these social metrics of behavioral function related to eye contact were linked to the underlying neurophysiology.

## Methods and Materials

### Participants

Participants included 17 healthy Autism Spectrum Disorder (ASD) adults (3 female; mean age 25±4.9 years; 12 right-handed, 3 left-handed, and 2 ambidextrous (51)) whose diagnoses were verified by gold standard, research-reliable clinician assessments, including the Autism Diagnostic Observation Schedule, 2^nd^ Edition (ADOS-2 (49)) (Table S1), and expert clinical judgment using DSM-5 criteria (6); and 19 healthy, typically-developed (TD) adults (mean age 26±5.8 years; 18 right-handed and 1 ambidextrous) (Table S2). Participants were recruited from ongoing research in the McPartland Lab, the Yale Developmental Disabilities Clinic, and the broader community through flyers and social media announcements. Inclusion criteria included age 18-45 years, IQ≥70, and English speaking. Exclusion criteria included diagnosis of bipolar disorder, personality disorder, or schizophrenia spectrum disorder; anti-epileptic, barbiturate, or benzodiazepine medication use; history of seizures, brain damage, or recent serious concussion; alcohol use within 24 hours; recreational drug use within 48 hours; chronic drug abuse; medication changes within two weeks; sensory impairment or tic disorder that would interfere with fNIRS recording; history of electroconvulsive therapy; or genetic or medical condition etiologically related to ASD. Additional exclusionary criteria for TD participants included self-report of any psychiatric diagnosis or learning/intellectual disability; psychotropic medication; or a first-degree relative with ASD. All participants provided written and verbal informed consent in accordance with guidelines and regulations approved by the Yale University Human Investigation Committee (HIC #1512016895) and were reimbursed for their participation. Assessment of the ASD participants’ capacity to give informed consent was provided by a consensus of trained professional staff who monitored the process and confirmed verbal and non-verbal responses. In order to assure that participants were comfortable during the experimental procedure, ASD participants were accompanied at all times by a member of the clinical team, who continuously evaluated their sustained consent to participate.

All participants were characterized by gender, age, full-scale IQ (FSIQ-4 as estimated by the Wechsler Abbreviated Scale of Intelligence, 2^nd^ Edition (WASI-II; (52), and self-reported clinical characteristics on several questionnaires, including the Autism-Spectrum Quotient (AQ; (53); Broad Autism Phenotype Questionnaire (BAPQ; (54); Social Responsiveness Scale, Second Edition (SRS-2; (50); Beck Anxiety Inventory (BAI; (55); State-Trait Anxiety Inventory (STAI; (56); and the Liebowitz Social Anxiety Scale (LSAS; (57)). See Tables S3 and S4 for detailed demographic and statistical comparisons between the two groups. Group comparisons of clinical assessments indicated expected differences on the AQ (p≤0.01); BAPQ (p≤0.01); SRS-2 (p≤0.01); and BAI scales (p≤0.05), and failed to provide evidence for differences on the WASI-II, STAI, and LSAS between the groups. Assessment and diagnostic tests were performed in clinical facilities at the Yale Child Study Center.

Participants were escorted from the clinical environment to the research environment for fNIRS / eye tracking experiments. An investigator was present during the data acquisition and monitored signs of discomfort during the experiment. All participants were paired with a same-gender TD lab partner. One male and one female, both in their 20’s, served as lab partners throughout the entire study. Lab partners were not informed of the participant’s group membership before the experiment.

Determination of a sample size sufficient for a conventional power of 0.80 was based on contrasts (Real face > Video Face) reported in a previous study using similar two-person techniques with a lab partner (47). Using the pwr package of R statistical computing software (58), a significance level of p < 0.05 is achieved with 16 participant/lab partner dyads. Sample sizes of 34 (17 ASD dyads) and 38 (19 TD dyads) ensure adequate effect sizes for these paired experiments. Although sufficient for the planned and reported analyses, a larger sample size would ordinarily be preferred. However, COVID related circumstances have prevented further acquisitions.

### Experimental Procedures and Paradigm

Dyads (participant and a gender-matched lab partner) were seated 140 cm across a table from each other and were fit with an extended head-coverage fNIRS cap. Each participant was either instructed to look straight ahead either at their partner or at a monitor with a video face adjusted in size to subtend the same visual angles as the real face (Fig 1A and B). In the live (Real Eye) task, dyads were instructed to gaze at each other’s eyes during cued 3-second epochs (1A), and in the video (Video Eye) task, dyads were instructed to gaze at the eyes of the face as it appeared in the dynamic video (1B). The illustrative red box enclosing the eyes of the participants in Fig 1 subtended 3.3x1.5° of visual angle and defined the location of the “eye box,” a region designated as the eye contact zone for each participant. In both tasks, dyads alternated their gaze between the eyes of their (real or video) partner and two small light-emitting diodes (LEDs) located 10° to the left and 10° to the right of their partner’s face (Figs 1C and D). The video was a recorded version of a same-gender participant performing the same task while wearing the same optode cap as live participants.

**Figure 1.**
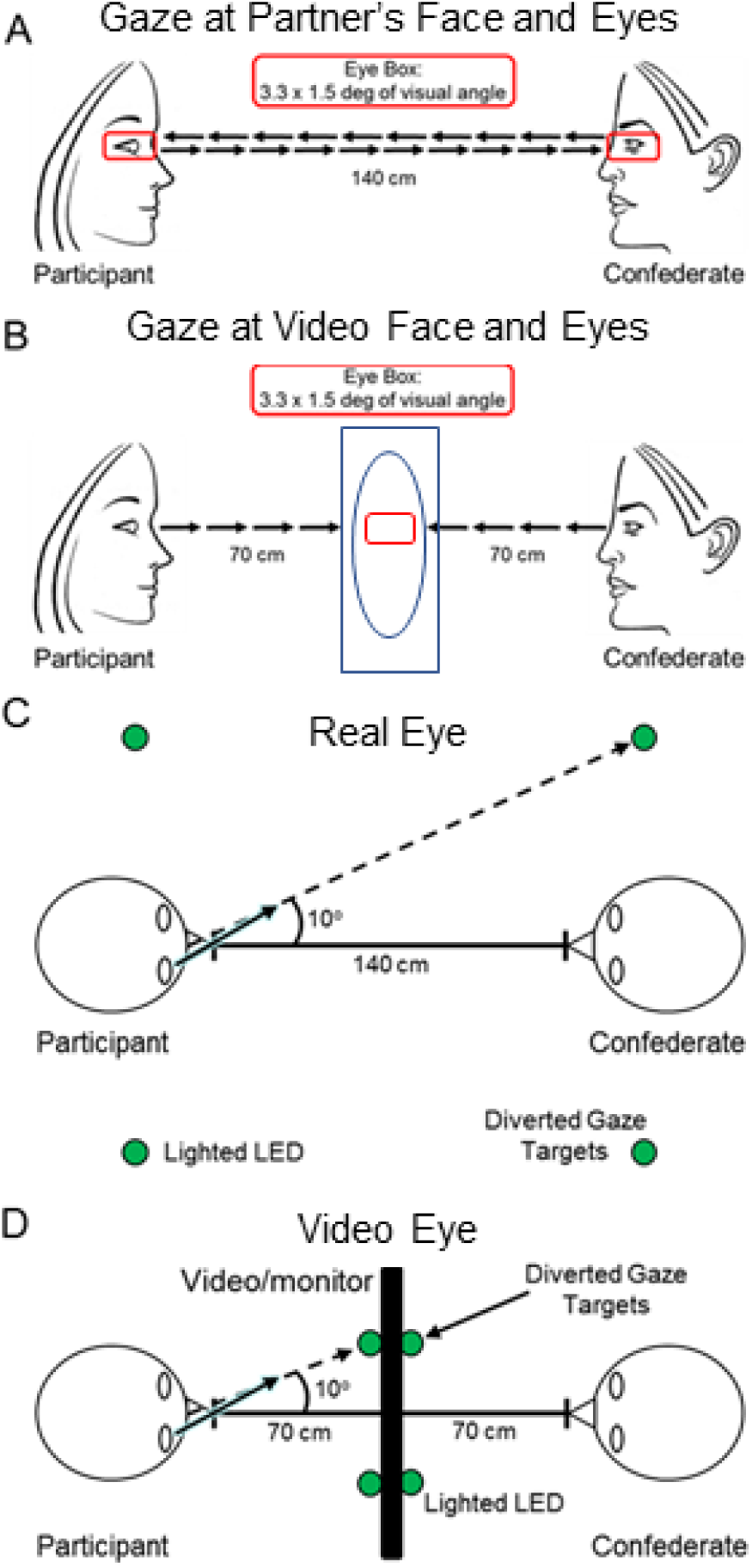
Eye gaze tasks. **A.** Gaze at partner’s eyes: Real Eye condition. Partners viewed each other at an eye-to-eye distance of 140 cm. The eye regions subtended by both the real eyes and the video eyes were 3.3 × 1.5 degrees of visual angle (red boxes). Small green LED indicator lights located to either side of their partner indicated rest and diverted gaze targets. **B.** Gaze at eyes in video: Video Eye condition. Two 24-inch 16x9 monitors were placed between the participants and a size-calibrated, pre-recorded dynamic video of a face was presented in the same field-of-view as the live interaction. **C.** Diagram of the Real Eye condition, with participant and lab partner sitting 140 cm apart from each other and LED indicator lights placed 10 degrees to the left and right of the Eye. **D.** Diagram of the Video Eye condition, with monitors arranged between partners. The face and LED sizes and positions were calibrated to subtend the same visual angles in both conditions.

The order of runs was randomly sequenced between viewing a real partner directly or viewing the visual-angle corrected video partner on a 24-inch 16x9 computer monitor placed back-to-back between participants, including a partition to assure that dyads could not see their real partner during video conditions. The face and distance of the video stimuli were calibrated to subtend identical degrees of visual angle in the field of view of the participants and the timing and range of motion of eye movements between partners were the same in both tasks. The time-series and experimental details were similar to prior studies (45, 47) and are included here to provide a self-contained report.

At the start of each task, an auditory cue prompted participants to gaze at the eyes of their real or recorded partner. Subsequent auditory tones alternatingly cued eye gaze between eyes or LED according to the protocol time series. The 15-second active task period alternated with a 15 s rest/baseline period. The task period consisted of three 6- second cycles in which gaze alternated on the partner for 3 s and then on a lighted LED to either the right or left (alternating) of the participant for 3 s for each of three events. The time series was performed in the same way for all runs. The order of runs was counterbalanced across pairs of participants. During the 15 s rest/baseline period, participants focused on the lighted LED, as in the case of the 3 s periods that separated the eye contact and gaze events. The 15 s activity epoch with alternating eye contact events was processed as a single block. The experimental paradigm (Fig 2A) employed a classic hemodynamic time series with 15 s of task alternating with 15 s of rest. Run length was 3 m and included six task-rest cycles. Due to the social discomfort associated with prolonged mutual gaze at another’s eyes, the task epochs were subdivided into events (epochs) that alternated between three 3-second “eye-on” and 3-second “eye-off” cycles (see Fig 2A). During the “eye-on” epoch, dyads were instructed to gaze at the eyes of their (real or video) partner, making eye contact as often as possible in natural intervals. An auditory tone signaled the transition between eye-on and eye-off events indicating when participants were instructed to divert their gaze to the LED targets 10° to the right or left.

**Figure 2.**
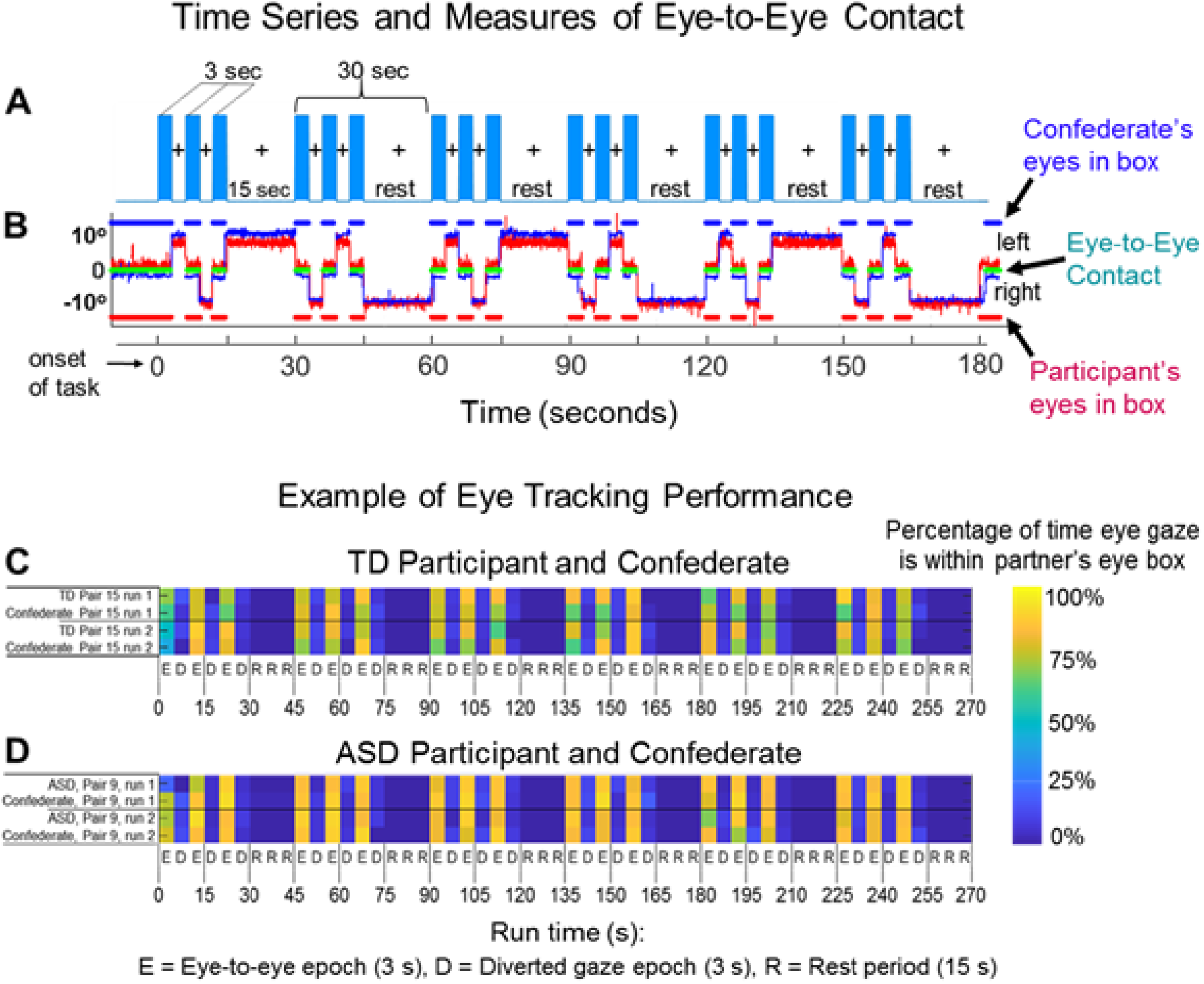
A. Time course. The duration of the run was three minutes and each run was repeated twice for both the Real Eye and Video Eye conditions. Each run included six alternating 15-second task and rest periods. In task periods (blue bars), participants alternated their gaze in three-second epochs between the eyes and the left or right lighted LED (See Fig 1, C and D). During the 15-second rest period, participants looked only at the lighted LED. The task is similar to those used in previous experiments (Hirsch et al., 2017; Noah et al., 2020). **B. Eye tracking traces of eye-to-eye contact.** Red traces represent eye movements from an ASD participant; blue traces represent the eye movements of a lab partner. The eye tracking data acquired on the Tobii system provides a frame-by-frame (8 ms) binary value that indicates whether or not eye gaze was directed within the eye-box of the partner. The blue dashed line (top) represents the duration of eye gaze (number of frames) that the lab partner’s gaze was within the eye-box of the participant. Similarly, the red dashed line (bottom) represents the duration of gaze (number of frames) that the participant’s eye gaze was in the eye-box of the lab partner. The green dashed line (middle) represents the length of time (number of frames) that the eyes of both partners were simultaneously focused within each other’s eye-boxes for a minimum of 83 ms. This is taken as a measure of eye-to-eye contact between the participant and the lab partner. **C, D. Gaze performance, Real Eye condition.** Eye contact epochs (3 s) are indicated as E; Diverted gaze epochs (3 s) are indicated as D; and Rest periods (15 s) are indicated as R. The color bar (top) indicates the percent of time eye gaze was within the eye box of the partner. **C.** Example eye tracking report for one TD participant and lab partner pair. **D.** Example eye tracking report for one ASD participant and the lab partner pair. Similar computations were performed for all participants when eye tracking data were acquired.

#### Eye Tracking

Two Tobii Pro x3-120 eye trackers (Tobii Pro, Stockholm, Sweden), one per participant, were used to acquire simultaneous eye tracking data at a sampling rate of 120 Hz. Eye trackers were mounted on the table facing each participant. Prior to the start of the experiment, a three-point calibration method was used to calibrate the eye tracker on each participant. The partner was instructed to stay still and look straight ahead while the participant was told to look first at the partner’s right eye, then left eye, then the tip of the chin. The same calibration procedure for video interactions was performed before recording on a still image presented on the monitor 70 cm in front of the participants. Similar live calibration procedures have been used successfully in prior investigations of in-person social attention (59, 60). As instructed for the eye movement task, participants alternated their gaze between ≈0° and 10° of deflection. Participants fixated on the eyes of the video (Video Eye condition) or the eyes of the lab partner (Real Eye condition) ±10° deflections to either the left or right. The eye contact portions of the task were 3 s in length, with six per trial, for 18 s of expected eye contact over the trial duration (Fig 2B).

Eye tracking provided a measure of compliance with task instructions to fixate on the eye as illustrated in Fig 2B for a lab partner (blue trace) and an ASD participant (red trace). The x-axis represents the run time series (180 s), and the y-axis represents the gaze angle, where 0 represents eye-to-eye contact and ±10° indicate left and right deflections, respectively. The moments of dyadic eye contact (gaze is within the eye box of their partner) are indicated by the green line. The time series of Fig 2A and 2B are synchronized for illustrative purposes. The blue and red dashed lines above and below the eye position trace indicate the times of gaze locations that are within the eye box of the partner for the lab partner and the ASD participant respectively. An “eye box hit” is defined when the gazes of both partners are within the designated eye box of the other for a minimum of 83 ms, i.e., 10 frames (61). The green lines in the figure indicate these 10-frame time points where the gaze of both partners was in the eye box of the other. The eye contact performance is illustrated for a typical participant in Fig 2C and an ASD participant in Fig 2D by the color of the event bar where the percentage of time in the eye box of the lab partner is represented by the color bar for the entire run time (180 s). Examples of average gaze positions are shown for 5 typical participants in Fig 3A to illustrate the gaze and eye box confirmation.

**Figure 3.**
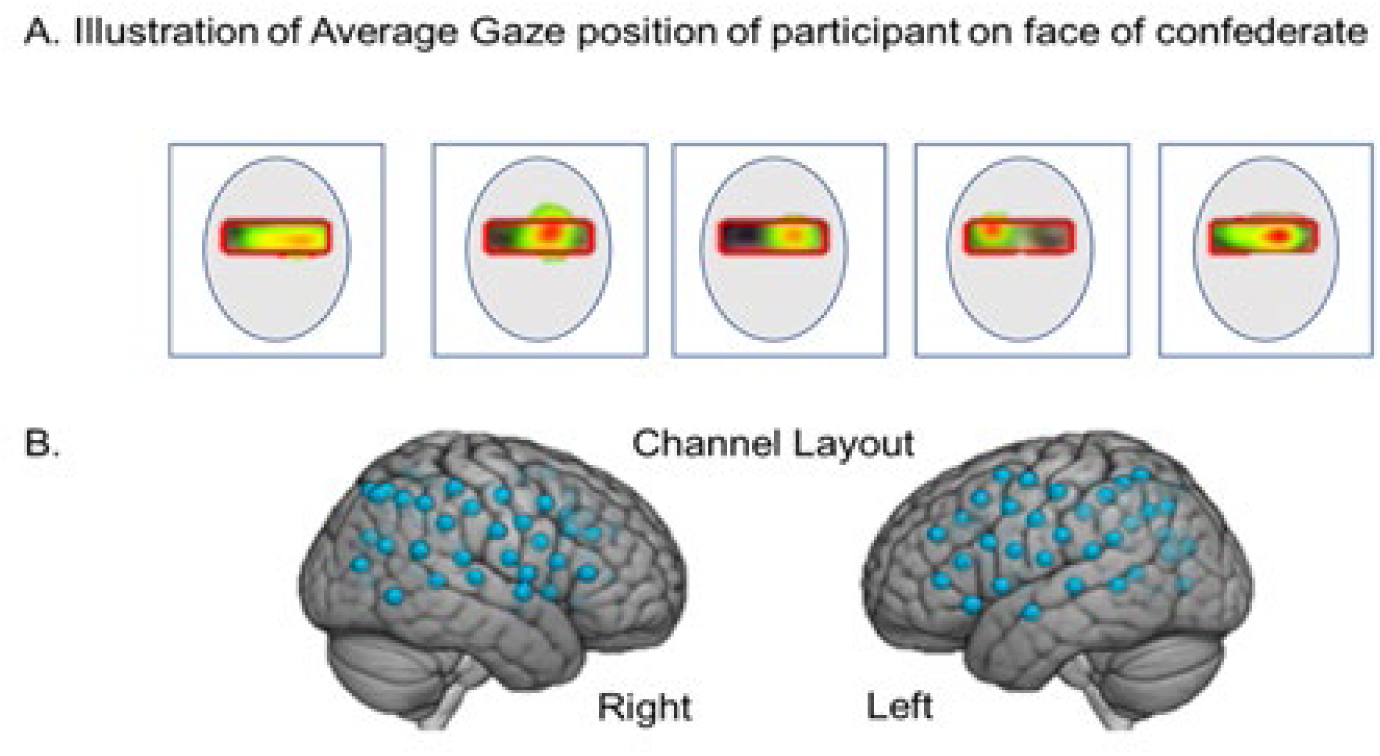
A. Examples of average participant eye gaze positions when viewing the face of the lab partner. The red box illustrates the target “eye box” and the color gradient from red to green indicates percent of “target hits” in the eye box for an entire run. **B.** fNIRS channel layout. Right and left hemispheres of a single rendered brain illustrate median locations (blue dots) for 58 channels per participant. Montreal Neurological Institute (MNI) coordinates were determined for each channel by digitizing emitter and detector locations in relation to anterior, posterior, dorsal, and lateral fiduciary markers based on the standard 10-20 system.

#### Functional NIRS Signal Acquisition and Channel Localization

Functional NIRS signal acquisition, optode localization, and signal processing, including global mean removal, were similar to methods described previously (62–68) and are briefly summarized below. Hemodynamic signals were acquired using 3 wavelengths of light, and an 80-fiber multichannel, continuous-wave fNIRS system (LABNIRS, Shimadzu Corp., Kyoto, Japan). Each participant was fitted with an optode cap with predefined channel distances. Three sizes of caps were used based on the circumference of the heads of participants (60 cm, 56.5 cm, or 54.5 cm). Optode distances of 3 cm were designed for the 60 cm cap but were scaled equally to smaller caps. A lighted fiber-optic probe (Daiso, Hiroshima, Japan) was used to remove all hair from the optode channel before optode placement.

Optodes consisting of 40 emitters and 40 detectors were arranged in a custom matrix, providing a total of 54 acquisition channels per participant. The specific layout with the coverage of the optode channels is shown in Fig 3B. For consistency, placement of the most anterior channel of the optode holder cap was centered 1 cm above nasion. To assure acceptable signal-to-noise ratios, resistance was measured for each channel prior to recording, and adjustments were made for each channel until all recording optodes were calibrated and able to sense known quantities of light from each laser wavelength (63, 69, 70). Anatomical locations of optodes in relation to standard head landmarks were determined for each participant using a Patriot 3D Digitizer (Polhemus, Colchester, VT) (71–75). Montreal Neurological Institute (MNI) coordinates (76) for each channel were obtained using NIRS-SPM software (77) with WFU PickAtlas (78, 79).

#### Analysis of eye-to-eye contact, dwell time, and pupillary responses

Eye tracking information including pupil size was exported from the Tobii system to the data processing pipeline and custom scripts in MATLAB where it was used to calculate the mutual eye contact events, accuracy, latency to targets, and pupil diameters. Eye-tracking data were not usable on 5 out of 17 ASD participants and 4 out of 19 TD participants due to either calibration or equipment problems (right columns of Tables S1 and S2 summarize the eye tracking acquisitions). Tobii Pro Lab software (Tobii Pro, Stockholm, Sweden) was used to create areas of interest for subsequent eye tracking analyses run in MATLAB 2014a (Mathworks, Natick, MA). The eye box was identified manually for each run and each participant for both live and video sequences. For the measures of gaze duration and variability, the horizontal components of gaze trajectories were gated by the eye-to-eye portions of each trial, retaining only samples that were within the eye box range.

This analysis used the zero angle (eye contact) intervals to characterize participant eye contact behavior. The eye tracking source was the horizontal component of post-processed trajectories converted to units of arc length (tenths of a degree). There were 1350 observations of 27 participants (15 TD, 12 ASD). To avoid possible inclusion of the large movements into and out of the valid range, the first and last 200 ms of each 3 s eye contact interval were excluded. Three measures were obtained from each interval: Dwell Time, the number of valid retained samples per interval normalized by sampling rate (seconds); Gaze Variability, the standard deviation of the samples centered over each interval, normalized by the number of retained samples (Figs 4A and B); and pupil diameter (mm) Figs 4C and D. In the case of the gaze position data (Figs 4A and B) linear mixed-effect models were used to assess the fixed effects of group (TD, ASD) and condition (Video Eye, Real Eye), with random intercepts by participant. Pupil sizes for left and right eyes (Figs 4C and D) were sampled at 40 Hz using the Tobii eye tracking system. All analyses used average pupil sizes across both eyes. To match the temporal resolution of the gaze position data, the pupil diameter data were interpolated to a sample rate of 120 Hz.

**Figure 4.**
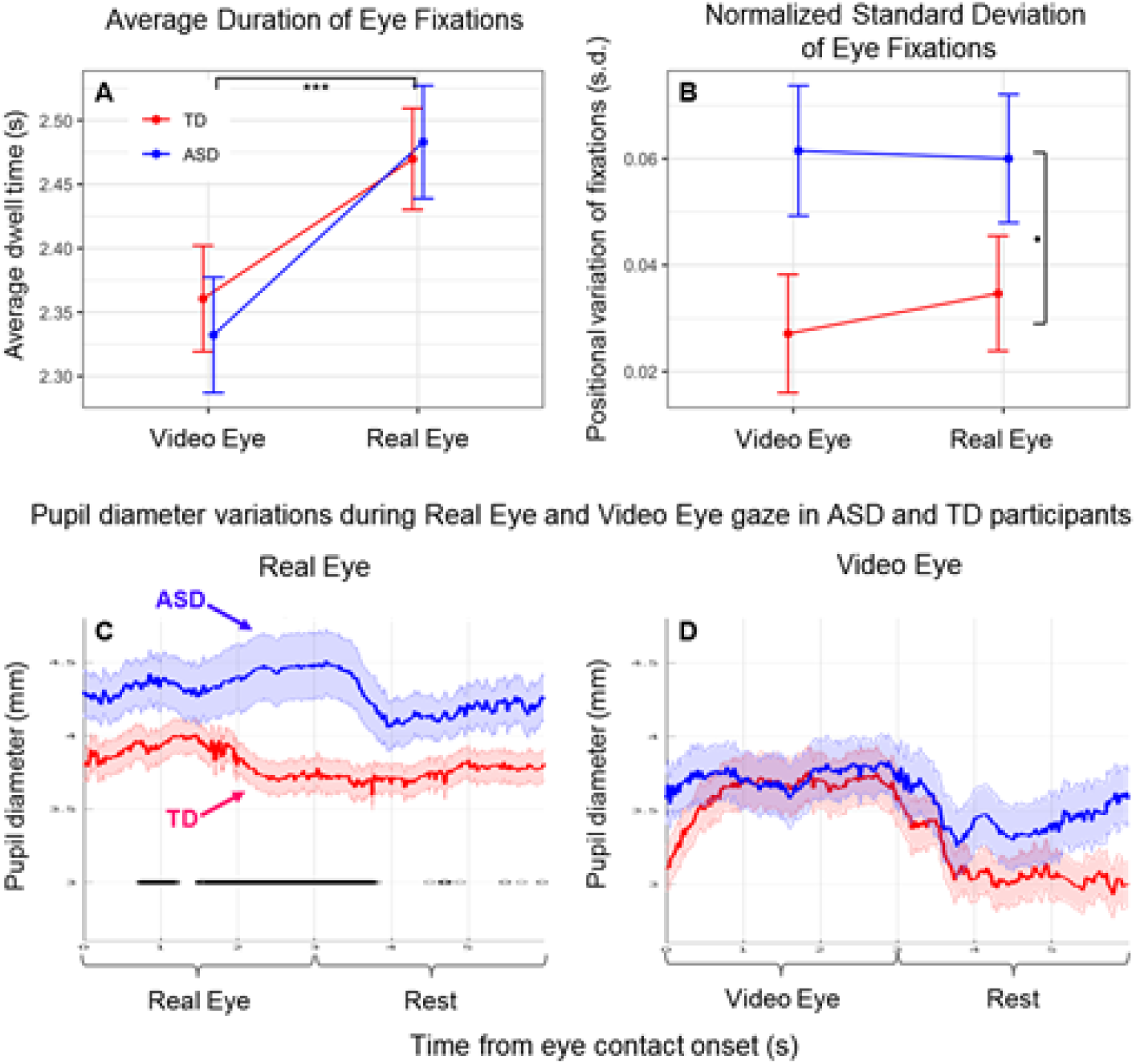
A, B. Marginal means plots based on linear mixed effects models for each measure by participant group. (Blue: Autism Spectrum Disorder (ASD) participants; Red: Typically- developed (TD) participants). **A.** Dwell time duration of eye contact on either the eyes in the video (Video Eye condition) or eyes of the lab partner (Real Eye condition). **B.** Standard deviation of horizontal gaze trajectory normalized by duration of contact. Error bars show SEM. ***p≤0.001, *p≤0.05. **C, D. Pupil diameter variations** observed in ASD and TD participants during Real Eye (**C**) and Video Eye (**D**) gaze conditions (Blue: Autism Spectrum Disorder (ASD) participants; Red: Typically-developed (TD) participants). Black lines indicate points at which pupil diameter differences between groups were significant at p≤0.05.

### fNIRS Signal Processing

Raw optical density variations were acquired at three wavelengths of light (780 nm, 805 nm, 830 nm), which were translated into relative chromophore concentrations using a Beer-Lambert equation (80–82). Signals were recorded at 30 Hz. Baseline drift was removed using wavelet detrending provided in NIRS-SPM (77). In accordance with recommendations for best practices using fNIRS data (83), global components attributable to blood pressure and other systemic effects (84) were removed using a principal component analysis (PCA) spatial global mean filter (62, 64, 85) prior to general linear model (GLM) analysis. All analyses are reported using the combined OxyHb and deOxyHb signals. The deOxyHb signal is inverted so that a positive result corresponds to increases in brain activity, similar to the OxyHb signal. The combined signal averages are taken as the input to the second level (group) analysis (86). Comparisons between conditions were based on GLM procedures using the NIRS-SPM software package. Event epochs within the time series were convolved with the hemodynamic response function provided from SPM8 (87) and were fit to the signals, providing individual “beta values” for each participant across conditions. Group results based on these beta values were rendered on a standard MNI brain template (TD-ICBM152 T1 MRI template (Mazziota et al., 2001) in SP8 using NIRS-SPM software (77) with WFU PickAtlas (78, 79).

### Code Accessibility

Custom code will be provided upon request at fmri.org.

## Results

### Behavioral

#### Eye-to-eye contact

Even though eye-to-eye contact is often reduced in individuals with ASD, in this investigation, we asked our participants to look directly at the face of the lab partner and to make eye-to-eye contacts during the cued 3-second periods. The recorded measures of gaze time in the “eye box” did not differ systematically between TD and ASD participants for either the Real Eye or Video Eye conditions (see Supplementary S1 and S2 Figs), confirming compliance with this task: a t-test of median eye box dwell time percentages failed to provide evidence for differences between groups (see Supplementary Table S5). This approach supports the assumption that both TD and ASD participants performed the same task, i.e., eye-to-eye contact during the 3 s epochs. Gaze dwell time assessed using a linear mixed effects model with fixed effects of Group and Condition and random intercepts by participant showed no group difference or interaction, but dwell time for both TD and ASD groups was significantly longer in the Real Eye condition (t=10.88, p≤0.001). See Fig 4A. However, gaze variability (assessed as the standard deviation of the horizontal component of eye trajectory during the eye contact intervals normalized by their duration) was greater for the ASD than the TD group for both conditions, see Fig 4B, consistent with altered visual sensing mechanisms in eye-movement patterns in ASD while viewing the face and eye stimuli in either condition (t=2.08, p≤0.05). Event-triggered averages of pupil diameter measurements were compared for the two conditions, real face-to-face (Fig 4C) and video face/eye gaze (Fig 4D). While both groups initially showed pupil dilation for gaze at real eyes, overall dilation in the ASD group was greater than in the TD group (p<0.05). No evidence for differences was observed between groups during gaze at the eyes in a video face suggesting increased autonomic responses to real faces, but not video faces, in ASD (88).

### Neural Findings

#### Direct comparison of TD and ASD findings for real eye-to-eye contact

A test of the alternative neural pathways hypothesis for live and interactive face processing in ASD is shown in Fig 5 with a direct comparison of the TD and ASD neural responses during live eye-to-eye contact. Red clusters indicate increased neural activity for the TD group and blue clusters indicate increased neural activity for the ASD (p≤0.05 FDR-corrected). Neural activity in the Real Eye condition is increased in the dorsal parietal regions for TD> ASD and in the temporal and ventral frontal regions for ASD>TD. See caption and tables for specific anatomical details.

**Figure 5.**
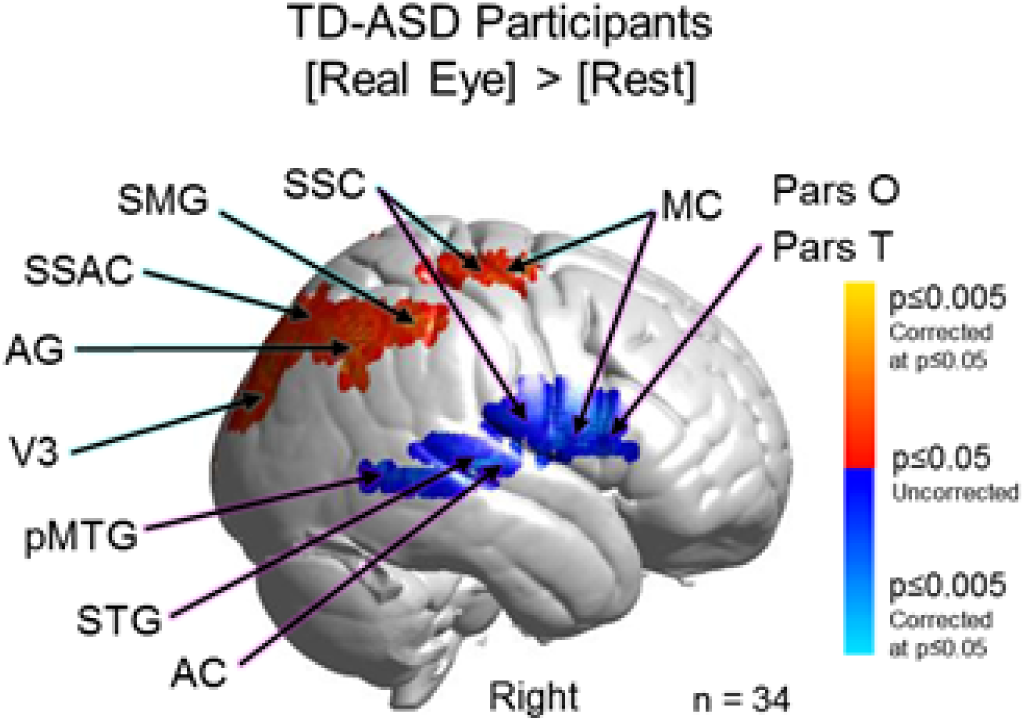
Contrast comparison [Real Eye > Rest], Typically-Developed (TD) participants relative to Autism Spectrum Disorder (ASD) participants. TD participants (red clusters) show comparatively greater activation in dorsal somatosensory cortex (SSC); supramarginal gyrus (SMG); angular gyrus (AG); pre- and supplementary motor cortex (MC); and extrastriate visual cortex (V3), while heightened activity was observed for ASD participants (blue clusters) in relatively ventral MC; SSC; pars opercularis (Pars O) and pars triangularis (Pars T); posterior middle temporal gyrus (pMTG); superior temporal gyrus (STG); and auditory cortex (AC). See Table 1. Yellow and light blue indicate responses corrected for multiple comparisons using FDR at p≤0.05. GLM analyses are based on the combined OxyHb and deOxyHb signals.

### Modulation of neural circuitry by frequency of eye contact events

A further test of the ventral vs dorsal alternative pathways hypothesis is shown in Fig 6. Neural responses during each 3 s eye viewing period were modulated by the number of eye contact events within that period for both TD and ASD groups. The covariance variable of eye-contacts used in the second level (group) analysis was constructed by assigning each subject with the median eye contact time for the 3 s periods where the eye of the partner was viewed. For TD participants (Fig 6A) clusters were observed in right dorsal supramarginal gyrus (SMG), somatosensory association cortex (SSAC), and dorsolateral prefrontal cortex (DLPFC); frontal eye fields (FEF); and pre- and supplementary motor cortex (MC) (see Table 2A). In sharp contrast to these TD observations, ASD participants’ (Fig 6B) neural responses to eye- to-eye signals modulated by the same measures of eye contact events were observed in the ventral right supramarginal gyrus (SMG); angular gyrus (AG); extrastriate visual (V3) and visual association cortices (V2); as well as the dorsolateral prefrontal cortex (DLPFC). Neural patterns in TD and ASD participants both demonstrated activity in the DLPFC whereas group response patterns were clearly differentiated in the posterior regions of the brain. In the case of TD participants, dorsal parietal regions were responsive to eye-to-eye contact, while in the ASD participants, ventral occipital and temporal regions were responsive to eye-to-eye contact. In summary, the findings are consistent with the hypothesis of alternative neural pathways for live eye contacts between TD and ASD groups. In particular, in TD the right dorsal parietal stream is activated, whereas in ASD, the right ventral occipital-temporal stream is activated.

**Table 1.**
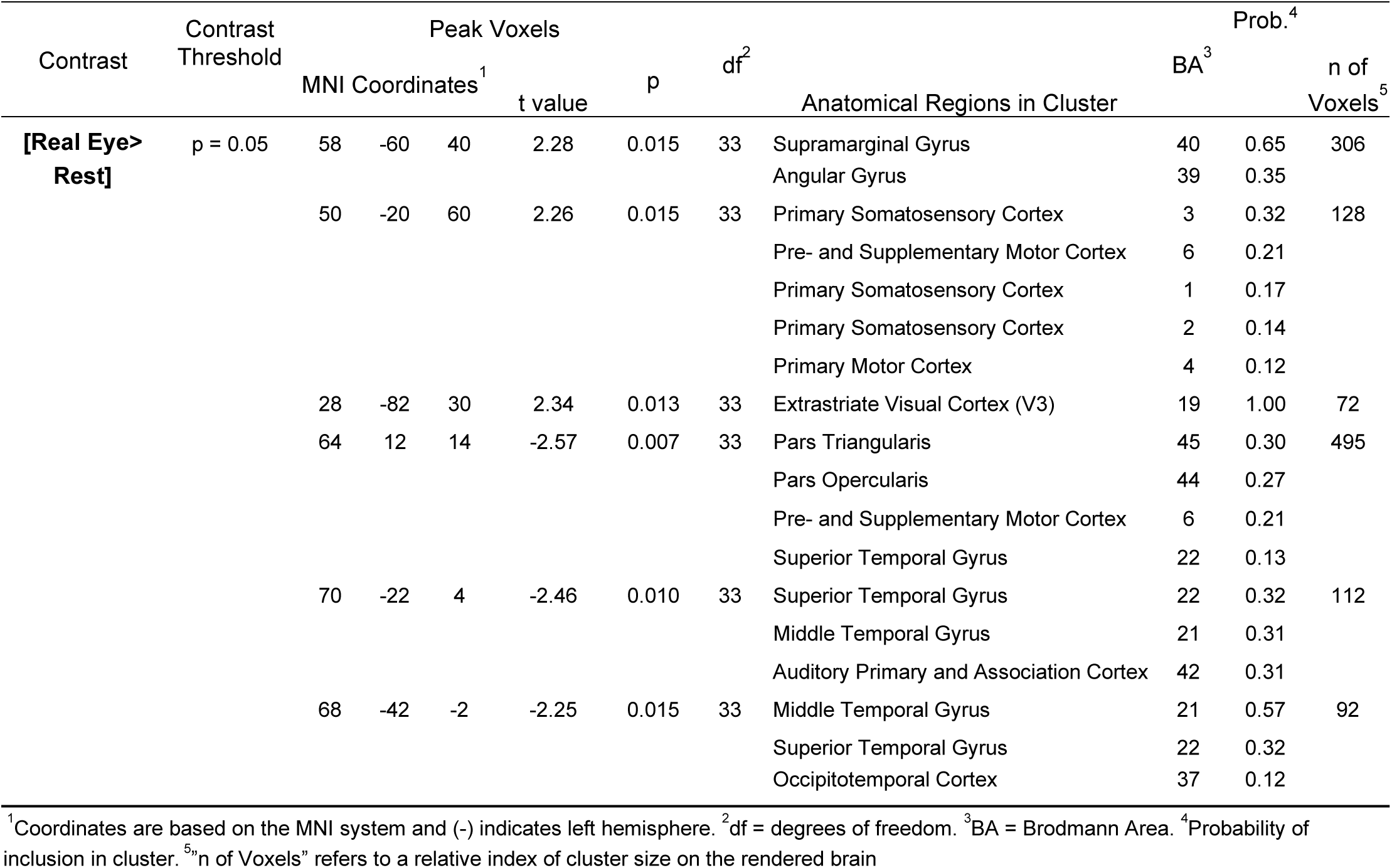
GLM Contrast comparison: [Real Eye] > [Rest] (deOxyHb + OxyHb signals), TD group - ASD group

**Table 2A.** GLM Contrast comparison: [Real Eye] > [Rest] with eye contact regressor (deOxyHb + OxyHb signals), TD group.

| Contrast |  | Peak Voxels |  |  |  |  |  |  |  |  |  |
| --- | --- | --- | --- | --- | --- | --- | --- | --- | --- | --- | --- |
| Contrast | Threshold | MNI Coordinates<sup>1</sup> |  |  | t value | p | df<sup>2</sup> | Anatomical Regions in Cluster | BA<sup>3</sup> | Prob. | n of Voxels |
| [Real Eye >Rest] | p = 0.05 | 58 | -54 | 48 | 3.78 | 0.0012 | 13 | Supramarginal Gyrus | 40 | 0.93 | 344 |
|  |  | 58 | 12 | 38 | 4.76 | 0.0002 | 13 | Dorsolateral Prefrontal Cortex | 9 | 0.52 | 342 |
|  |  |  |  |  |  |  |  | Pre- and Supplementary Motor Cortex | 6 | 0.30 |  |
|  |  |  |  |  |  |  |  | Frontal Eye Fields | 8 | 0.16 |  |
|  |  | 26 | -62 | 54 | 2.37 | 0.0170 | 13 | Somatosensory Association Cortex | 7 | 1.00 | 18 |
|  |  | 62 | -2 | 42 | 3.06 | 0.0045 | 13 | Pre- and Supplementary Motor Cortex | 6 | 0.82 | 10 |

<sup>1</sup>Coordinates are based on the MNI system and (-) indicates left hemisphere. <sup>2</sup>df = degrees of freedom. <sup>3</sup>BA = Brodmann Area.

**Table 2B.**
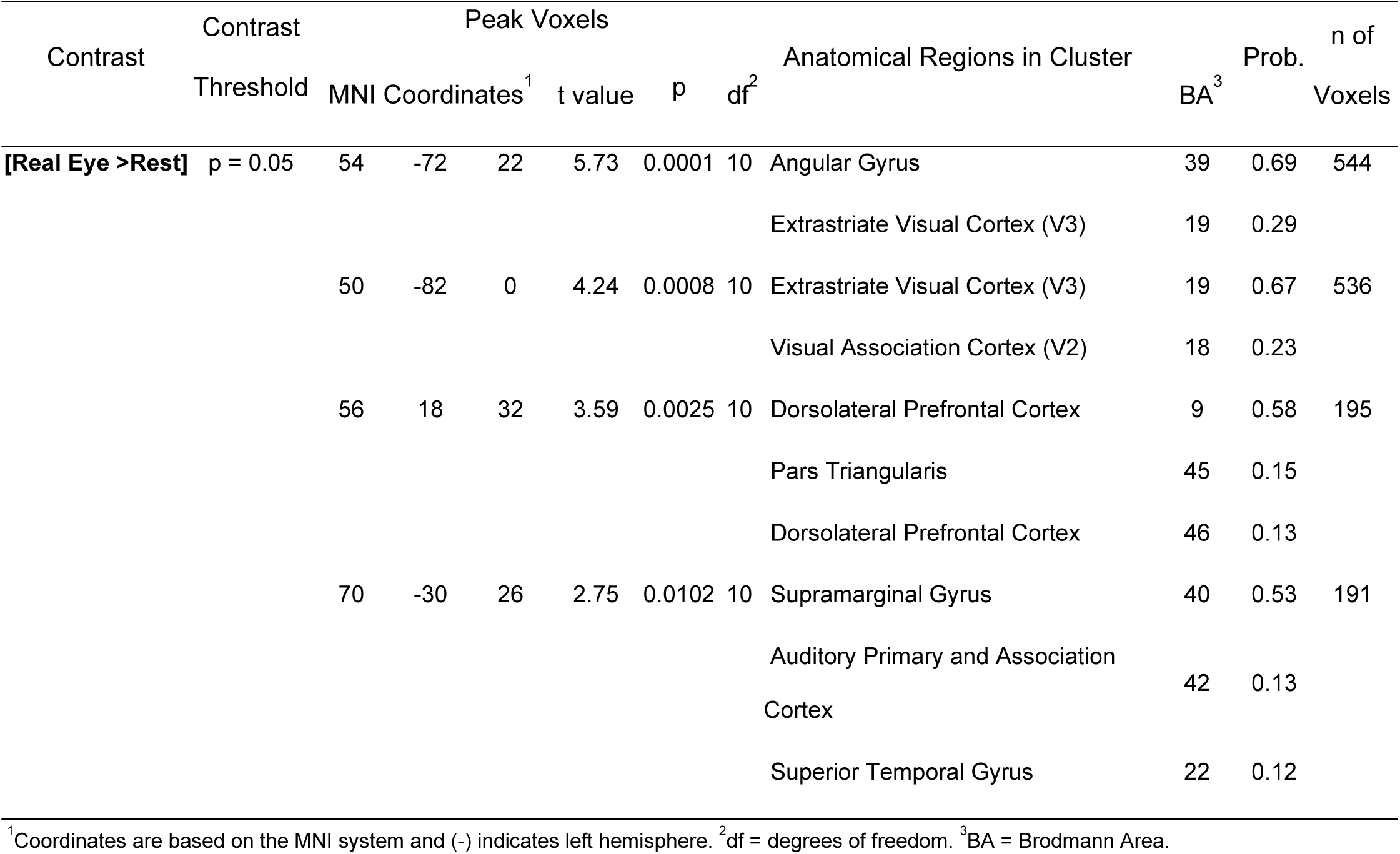
GLM Contrast comparison: [Real Eye] > [Rest] with eye contact regressor (deOxyHb + OxyHb signals), ASD group.

**Figure 6.**
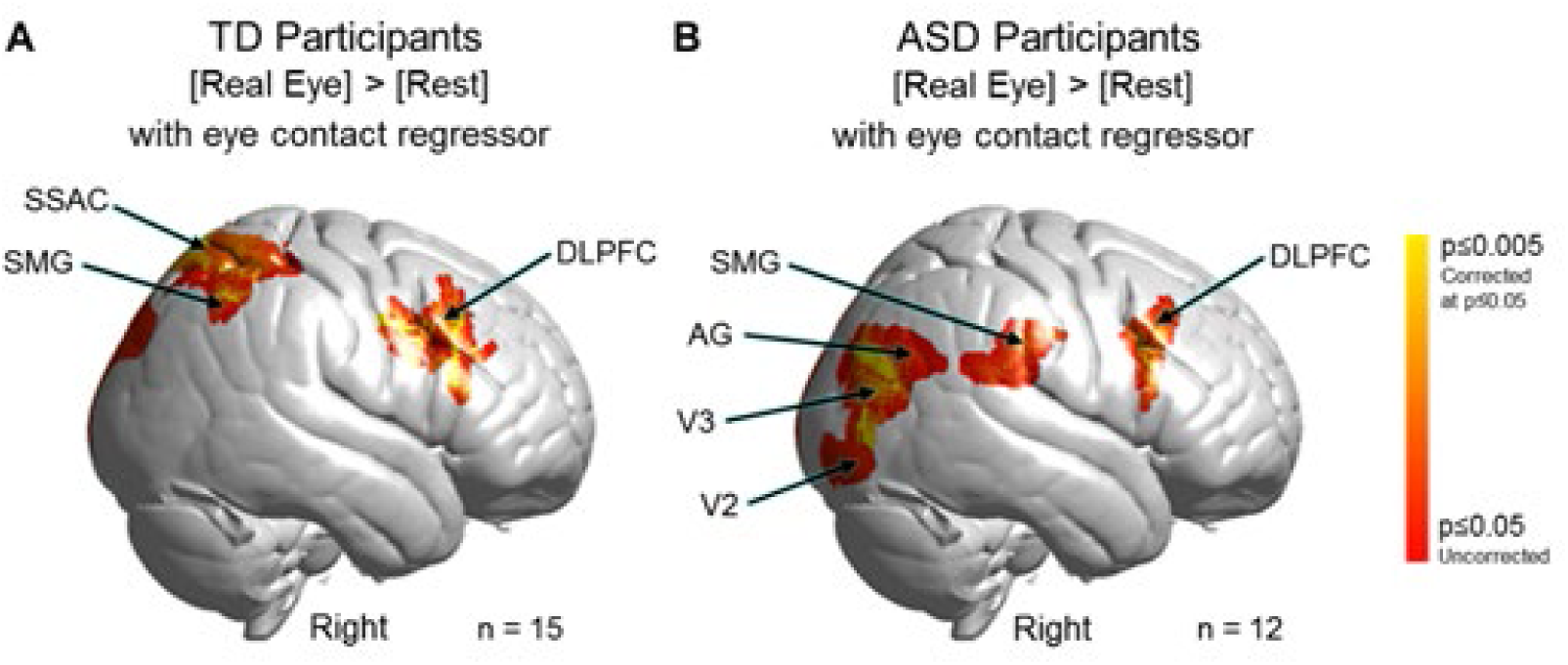
Contrast comparison [Real Eye] > [Rest] modulated by the number of frames within each 3-second eye viewing period where the gaze of both partners was simultaneously within the eye box of the other. A. Typically-developed (TD) participants. Activity observed in the right hemisphere: supramarginal gyrus (SMG); somatosensory association cortex (SSAC); dorsolateral prefrontal cortex (DLPFC); frontal eye fields (FEF); and pre- and supplementary motor cortex (MC). See Table 2A. Note: n = 15 rather than 19 (see Table S2) because eye tracking data could not be acquired on four participants. B. Autism Spectrum Disorder (ASD) participants. Activity observed in the right hemisphere include SMG; angular gyrus (AG); extrastriate visual cortex (V3); visual association cortex (V2); and DLPFC. See Table 2B. Note: n = 12 rather than 17 (see Table S1) because eye tracking data could not be acquired on five participants. Yellow indicates signals corrected for multiple comparisons at p≤0.05 using FDR. GLM analyses are based on the combined OxyHb and deOxyHb signals.

#### Neural responses during real eye gaze modulated by symptom severity as measured by ADOS-2: Group effects

Neural responses (beta values) acquired during eye-gaze were regressed by the individual ADOS-2 scores using the general linear model, GLM. The whole- brain main effect of the eye contact activity modulated by ADOS-2 scores is shown in Fig 7. Blue clusters indicate regions of the brain where neural activity as represented by the individual average was negatively related to the individual ADOS-2 scores. That is, participants with higher ADOS-2 scores and greater symptomatology showed consistently lower live eye contact related neural activity located in the right dorsal parietal areas including angular gyrus (AG), supramarginal gyrus (SMG), somatosensory association cortex (SSAC), and somatosensory cortex (SSC). Similar findings were also observed the right dorsolateral prefrontal cortex (DLPFC) (p≤0.01). See Table 3.

**Table 3.**
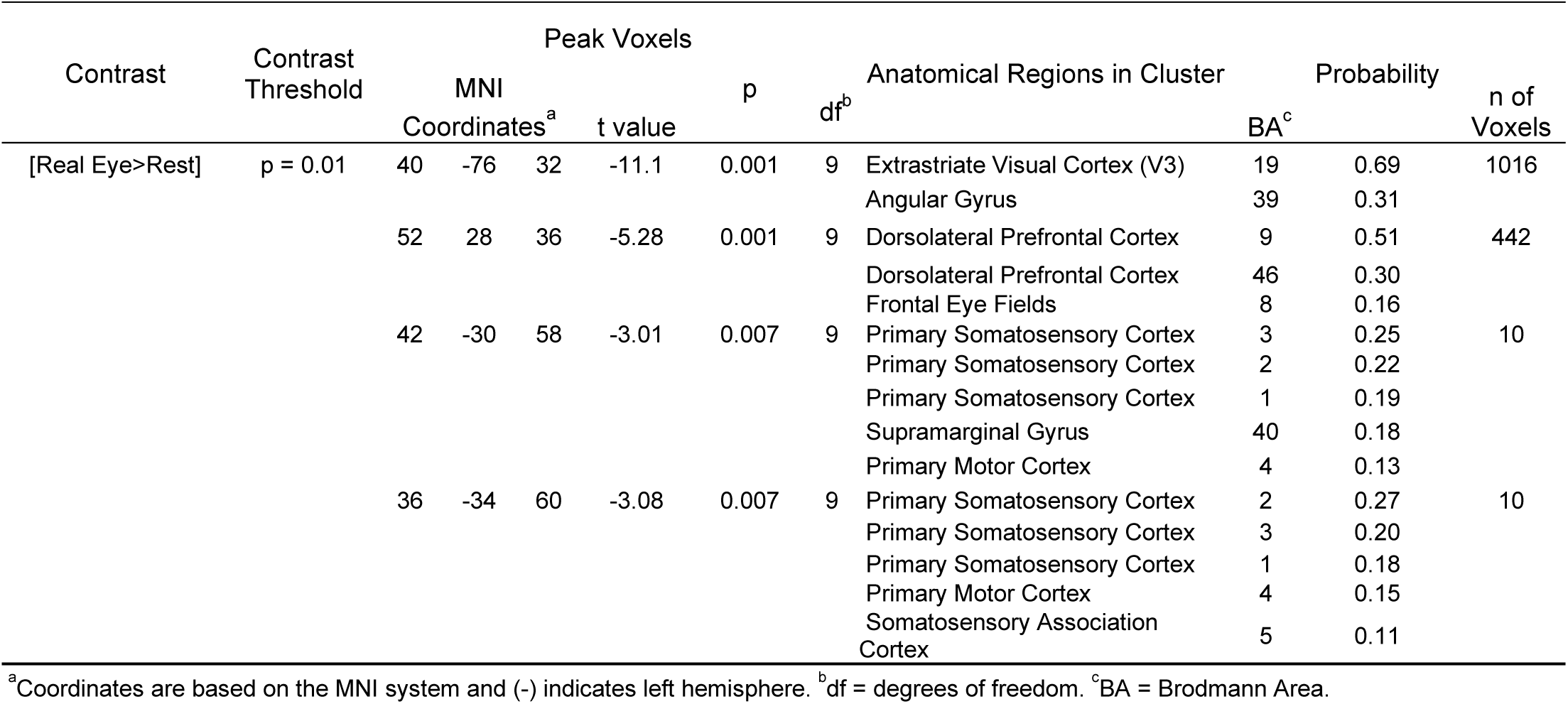
GLM Contrast comparison: [Real Eye >Rest] with ADOS-2 regressor (deOxyHb + OxyHb signals)

**Figure 7.**
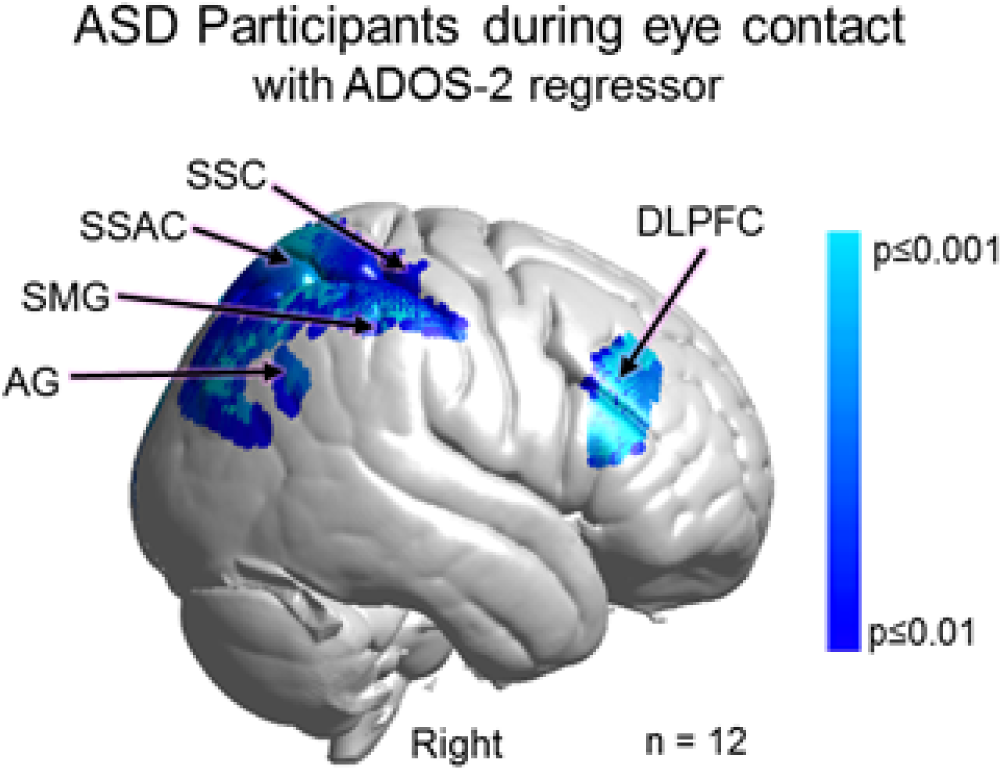
Main effect neural results for Autism Spectrum Disorder (ASD) participants during real eye contact with ADOS-2 (Autism Diagnostic Observation Schedule, 2^nd^ Edition) scores as a linear regressor. OxyHb and deOxyHb signals are combined. See Table 3. Blue colors indicate a negative relationship between neural responses and ADOS scores indicating that as symptom severity increases, neural responsiveness in these regions decreases. Light blue indicates responses corrected for multiple comparisons using FDR at p≤0.01. SSC: somatosensory cortex; SSAC: somatosensory association cortex; SMG: supramarginal gyrus; and AG: angular gyrus.

#### Neural responses during real eye gaze correlated with symptom severity as measured by ADOS-2: Individual ASD effects

The individual ADOS-2 scores for each participant (identified by participant number in Table S1) (x-axis) are plotted against the median beta- values, parameter estimations, of the fNIRS signal (y-axis) for the two regions of interest identified by the group effects above: A posterior dorsal stream cluster consisting of the somatosensory association cortex, supramarginal gyrus, angular gyrus, and somatosensory cortex, and an anterior cluster in the dorsolateral prefrontal cortex. The best fit lines illustrate the negative relationships for the posterior regions (r = -0.69, Fig 8A) and anterior frontal area (r = -0.77, Fig 8B).

**Figure 8.**
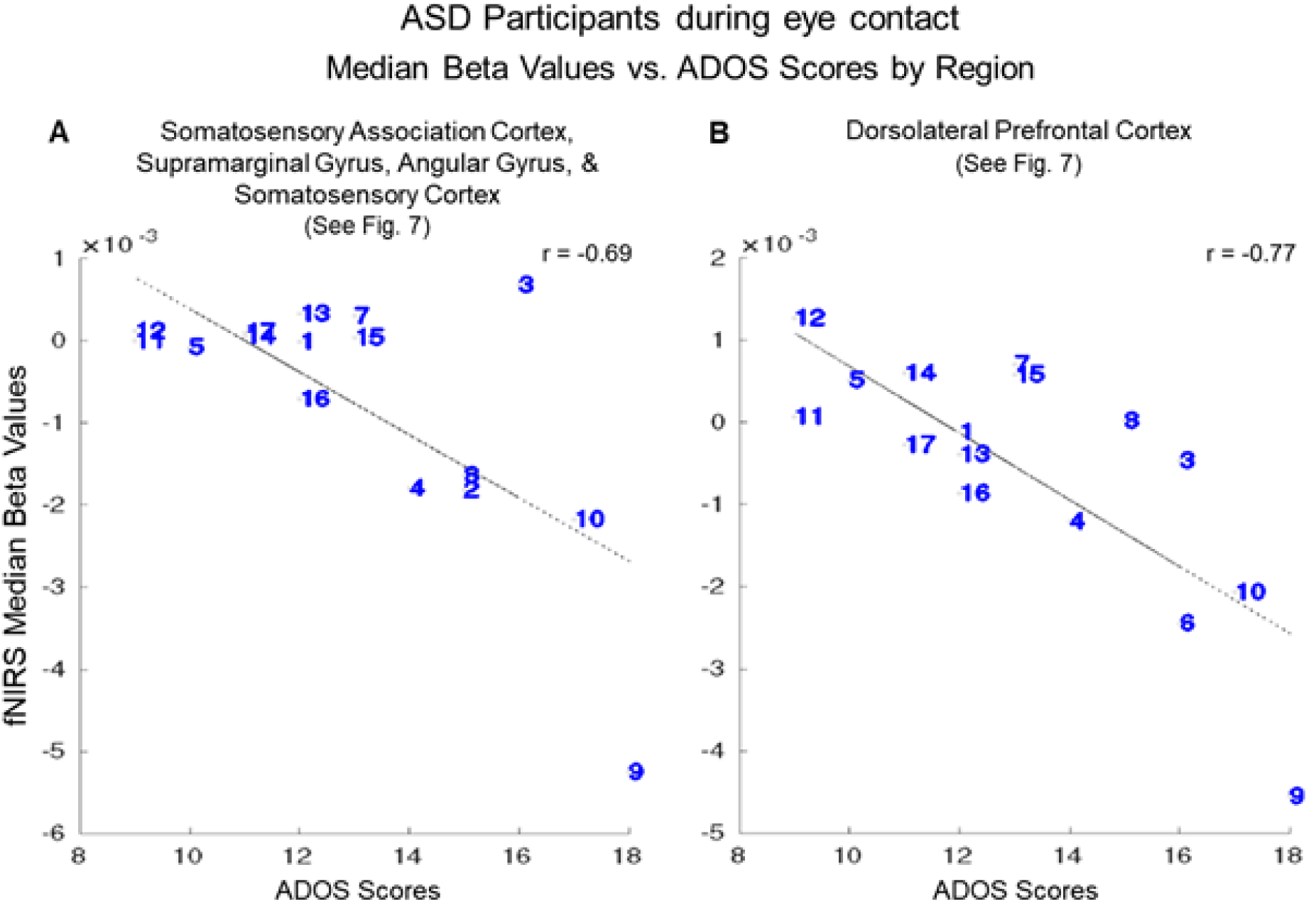
Autism Spectrum Disorder (ASD) participants (numbers correspond to Table S1) during eye contact beta values vs. ADOS-2 (Autism Diagnostic Observation Schedule, 2^nd^ Edition) scores. The median hemodynamic signals (Beta values, y-axis) within the responsive brain regions (Fig 7 and Table 3) and ADOS-2 scores (x-axis) are shown for each participant. The main effect of eye-to- eye contact is negatively correlated with fNIRS signals in **A.** right hemisphere somatosensory association cortex, supramarginal gyrus, angular gyrus, and somatosensory cortex (r = -0.69); and **B.** right dorsolateral prefrontal cortex (r = -0.77).

#### Neural responses during real eye gaze correlated with symptom severity as measured by SRS-2: Individual ASD and TD Effects

To further evaluate the relationship between social symptomatology and live-face eye-gaze for ASD and TD participants we combine the SRS-2 scores for both groups based on the assumption that ASD traits are also present in the general population. Consistent with the findings based on the ADOS-2 scores above, a negative relationship (blue cluster in Fig 9) was observed in regions located in the right dorsal stream including somatosensory cortex (SSC), somatosensory association cortex (SSAC), and supramarginal gyrus (SMG). Participants with higher SRS-2 scores indicating higher levels of symptomatology showed reduced neural activity during eye contact in the right somatosensory cortex (SSC), somatosensory association cortex (SSAC), and supramarginal gyrus (SMG) (p≤0.01). Table 4 provides the peak MNI coordinates, cluster t-values, anatomical regions within the cluster, Brodmann’s Area (BA), probability of inclusion in the cluster, and relative size of the active area (n of voxels).

**Figure 9.**
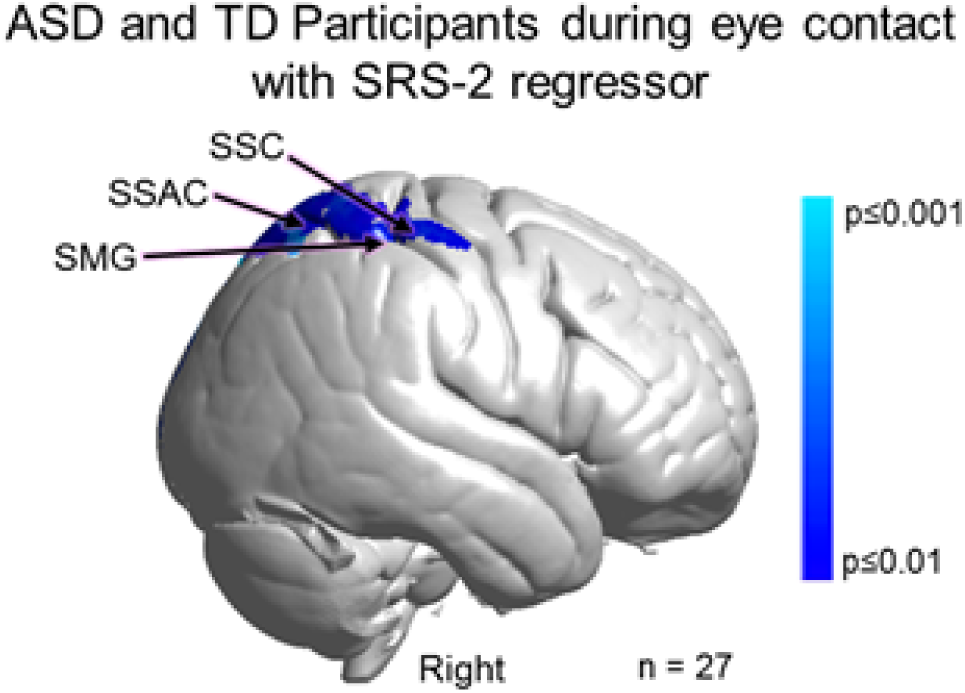
Main effect neural results for Autism Spectrum Disorder (ASD) and typically-developed (TD) participants during eye contact with SRS-2 (Social Responsiveness Scale, Second Edition) scores as a linear regressor. OxyHb and deOxyHb signals are combined. Blue colors indicate a negative relationship between neural responses and SRS-2 scores, which suggests that increased symptom severity is associated with reduced regional neural responsiveness (See Table 4). Light blue indicates responses corrected for multiple comparisons using FDR at p≤0.01. SSC: somatosensory cortex, SSAC: somatosensory association cortex, and SMG: supramarginal gyrus.

**Table 4.**
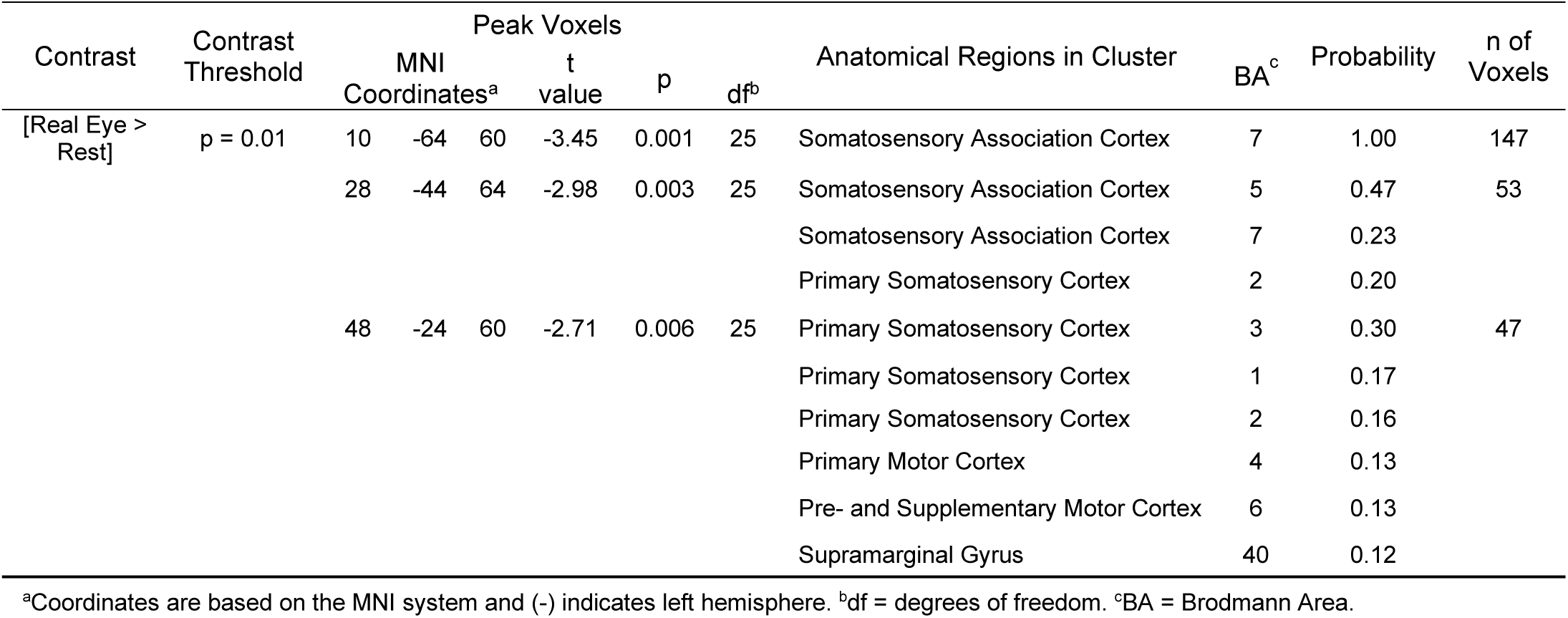
GLM Contrast comparison: [Real Eye>Rest] with SRS-2 regressor (deOxyHb + OxyHb signals)

#### SRS-2 scores and neural responses: Individual Differences

The individual SRS-2 scores for each participant (identified by a number that corresponds to the participant number in Tables S1 and S2) are plotted against the individual median beta values of the fNIRS signal (Fig 10). Red numbers represent TD participants and blue numbers represent ASD participants. The interspersal of the individual scores between ASD and TD participants is consistent with the assumption that social responsiveness traits vary within the general population as well as within ASD. The best fit line illustrates the negative relationship (r = - 0.58).

**Figure 10.**
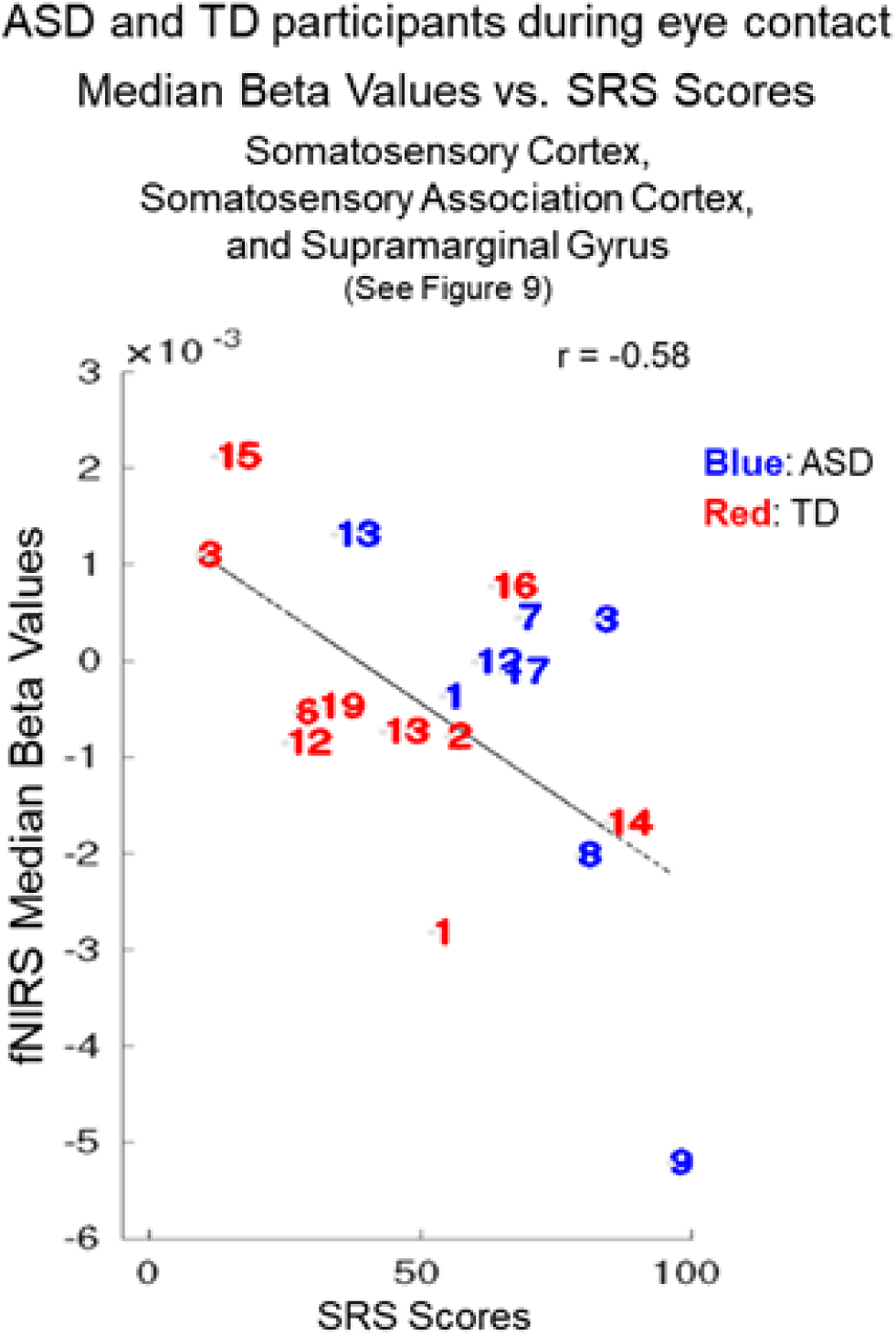
Autism Spectrum Disorder (ASD) participants (blue numbers) and typically-developed (TD) participants (red numbers) during eye contact vs. Social Responsiveness Scale (Second Edition, SRS-2) scores. The median hemodynamic signals (Beta values, y-axis) within the responsive brain region (Fig 9 and Table 4) and SRS-2 scores (x-axis) are shown for each participant. The main effect of eye-to-eye contact is negatively correlated with fNIRS signals in right hemisphere somatosensory cortex, somatosensory association cortex, and dorsal supramarginal gyrus (r = -0.58). Numbers indicate individual participants shown in Table S1 (ASD participants) and Table S2 (TD participants).

## Discussion

### Summary of findings

Overall, there were three main findings of this investigation: 1) the alternative neural processes hypothesis was supported for live and interactive faces in ASD.

Specifically, neural systems within ventral occipital-temporal regions were engage d in ASD whereas in TD these functions were associated with dorsal parietal and lateral prefrontal regions; 2) Variations in oculomotor and visual sensing were observed in ASD including increased positional variation in eye fixations and increased pupillary reactions to live faces, suggesting that visual acquisition factors may also contribute to live face processing mechanisms; and 3) A biological basis for social performance associated with ASD is suggested by the association between ADOS-2 and SRS-2 scores and the counter- correlation of neural responses in the right dorsal parietal regions during real eye-to-eye contact.

Differences in social performance, including reduced eye contact, are common characteristics of ASD. Although disparities in face processing and oculomotor variations in ASD are well documented, it is not known how these behaviors are linked to the underlying neurophysiology associated with live and natural interactions. In this investigation, we employed a two-person paradigm using functional near-infrared spectroscopy (fNIRS) to acquire neuroimaging responses during live dynamic eye-to-eye contacts with a lab partner that are a close proxy to the behaviors under investigation. Simultaneous eye-tracking and oculomotor responses were also acquired in 17 adult ASD dyads and 19 closely matched TD adult dyads. Social performance was quantified by clinical interview (ADOS-2: Autism Diagnostic Observation Schedule, 2^nd^ Edition) in ASD and self-report (SRS-2: Social Responsiveness Scale, Second Edition) in both ASD and TD to test the hypothesis that individual differences in social function are predicted by neural responses associated with live eye-to-eye contact.

Direct comparisons of neural findings between TD and ASD for the Real Eye condition are consistent with right dorsal parietal activity in the case of TD (TD > ASD) and right ventral occipital-temporal activity in the case of ASD (ASD > TD). Neural findings modulated by real eye-to-eye contact behavior revealed similar findings of increased right dorsal parietal activity for TD and alternatively increased right ventral parietal activity for ASD consistent with the hypothesis of dorsal eye processing systems for TD and ventral eye processing streams for ASD groups. Individual ADOS-2 scores were negatively correlated (r = -0.69) with individual fNIRS beta values (representing the strength of hemodynamic signals) within clusters in the right dorsal parietal stream including somatosensory cortices, angular gyrus, and supramarginal gyrus. Similarly, SRS-2 scores for the combined ASD and TD groups were negatively correlated (r = -0.58) with somatosensory cortices and the supramarginal gyrus also located in the right dorsal parietal stream. Since these two clinical measures provide similar information, it is expected that their relationship to underlying neural correlates would be similar. Neural responses in the right dorsolateral prefrontal cortex (DLPFC) during live eye-to-eye contact were also negatively correlated (r = -0.77) with ADOS-2 scores. These correlations between social function and the neural responses during live eye-to-eye contact are consistent with a model of dorsal parietal and dorsal lateral prefrontal cortex contributing to behavioral differences in ASD.

### Two-person Visual Sensing in ASD

Pupillometry, a presumed measure of activity associated with the locus coeruleus-norepinephrine system (88) revealed increased pupil diameters for ASD during real eye conditions but not the video eye conditions relative to TD (p<0.05), consistent with the interpretation of increased arousal associated with the real face and eyes. Further, variance of gaze positions was greater for ASD than TD for both real and video faces suggesting oculomotor differences in visual sensing (89).

A natural in-person encounter typically involves active visual sensing of dynamic face landmarks (90). Guidance systems for visual saccades and fixations are thought to actively “seek” relevant visual information such as social cues that are conveyed hierarchically to higher levels of neural processing (89). The cascade of synchronized oculomotor behaviors, for example, associated with mutual live eye-to-eye contacts does not occur during passive gaze at an inanimate representation of a real person because dynamic behaviors from both partners are required for a mutual eye contact event or a dynamic face-to-face interaction. In this investigation, we include measures of visual sensing, dwell time, and positional variance as well as pupil size to test the hypothesis that in ASD the live two-person condition may be distinguished by oculomotor behaviors in addition to neural processing. Differences in oculomotor functions have been previously reported in ASD, for example, for static and simulated stimuli (91, 92). Here we test the hypothesis that oculomotor systems are also affected during two-person face-to-face interactions.

The observed increased positional variation in ASD eye movements leads to the speculation that information characterizing an interactive face may not have been sufficiently acquired for ASD participants. The finding that live face processing in the ASD group increased activity in ventral and lateral occipital and temporal regions, rather than dorsal parietal regions, could be due, in part, to differences in visual sensing. The observed ventral processes are more consistent with non-interactive face functions than with interactive functions. For example, topographical maps associated with regional specializations for coding simulated faces are well-established. The ventral-occipital cortex is highly selective for and sensitive to pictures of faces (93, 94). Regions within the superior temporal sulcus are involved in detecting dynamic facial movements presented in two-dimensional stimuli (27, 29), and parameterized face processing codes for static faces have been identified by electrophysiology in middle and superior temporal gyri of non-human primates (95).

### Live Two-Person Interactions in ASD

Neuroimaging based on functional near-infrared spectroscopy (fNIRS) enables simultaneous acquisition of hemodynamic brain signals from two individuals (hyperscanning) dynamically engaged in natural interactions. Eye tracking acquired simultaneously on both participants during face-to-face engagement enables identification of eye contact events that occur between the partners. The aim to understand the neural mechanisms that underlie social function in ASD has motivated this multi-modal application of fNIRS and eye tracking. In spite of the biological significance of live interpersonal interactions for survival and social well-being, the underlying neural processes of interactive behaviors are relatively novel targets of investigation for natural settings as well as clinical, developmental, and psychiatric applications (37–39, 96). Fundamental models of dynamic and reciprocal behaviors are under development for multiple sensory and communication systems, clinical applications, and social behaviors (37-39, 97-102).

Current models of face and eye processing in TD and ASD are based primarily on non-interactive paradigms where data are acquired in single-subject situations using conventional stimulus and response models rather than dyadic paradigms that include live social interactions. The importance of investigations that include natural and dynamic two-person interactions between individuals is highlighted by a general theoretical framework proposed by the Interactive Brain Hypothesis (103, 104), which suggests that live interactions between individuals engage neural functions not activated during similar tasks performed alone, i.e., without interaction. A rapidly emerging neuroimaging literature and theoretical framework of live and natural interactions compared to single-subject interactions contributes an accumulating body of evidence in support of this hypothesis (31, 37, 39, 40, 67, 102, 105). Understanding neural activity during natural interactions is especially critical in ASD, as the defining social and communicative characteristics of the condition are often attenuated or absent during explicit laboratory tasks (41).

This long-standing experimental paucity of two-person interactive experimental paradigms in ASD, in part, reflects the historical limitations of conventional neuroimaging methods. For example, in functional magnetic resonance imaging (fMRI) solitary confinement in the bore of a scanner with minimal tolerance of head movements constrains/contraindicates investigations of natural, two-person interactions. Fortunately, however, these particular limitations are substantially resolved by recent developments of optical neuroimaging, functional near-infrared spectroscopy (fNIRS), a non-invasive spectral absorbance technique that detects changes in blood oxygen levels in both oxyhemoglobin and deoxyhemoglobin using surface-mounted optical sensors (106–109). Functional NIRS enables simultaneous acquisitions of hemodynamic signals (assumed to be a proxy for neural activity as in fMRI) from naturally interacting dyads and provides simultaneous dyadic measures that contribute to understanding interactive behaviors as opposed to single- subject responses that focus primarily on perceptual and cognitive systems.

### Social performance and face processing

This application of two-person neuroimaging technology to investigate the relationship between the neural underpinnings of interactive face and eye contact and social performance in ASD addresses a prominent and understudied question. Individual clinical evaluations of social performance applied as a second level regressor on whole-brain neuroimaging findings acquired during live real person eye-to-eye contacts confirm a negative relationship between test scores and neural signals in brain regions responsive to real eye-to-eye contacts. Participants with higher ADOS-2 scores, reflecting reduced social performance, showed lower neural signals (beta values, an indicator of signal strength and fit to the general linear model) in brain regions previously associated with social activity, interactive face processing, and motion sensitivity. Findings also included the right dorsolateral prefrontal cortex, a region implicated in both ASD and commonly co-occurring conditions, such as major depressive disorder. Further, a similar finding was observed for the SRS-2 when the scores of both TD and ASD participants were combined for regions within the dorsal stream but not the DLPFC. That is, as individual social ability decreased as indicated by the elevated SRS-2, the neural signal decreased in the right dorsal-stream regions. A similar finding was observed for SRS-2 scores and the relationship to neural signals acquired by fMRI in the fusiform gyrus and the amygdala during static face processing (110). Interestingly, in this study, the SRS-2 finding included TD as well as ASD participants suggesting that variations in social responsiveness and the associated reduction in dorsal stream activity are similarly represented in the general population.

Unique features of this study include the live interactive eye-to-eye task as well as the eye-tracking documentation of compliance combined with the individual difference approach to characterize single participants by both measures. All data were acquired during the epochs when participants directed their eye gaze to within the eye-box of the lab partner.

Continuous monitoring confirmed high levels of compliance in all cases. That is, when asked to perform the task, participants were able to do it although eye-to-eye contact was not necessarily a comfortable task for them.

These findings of a negative association between right dorsal regions and social performance do not imply a causal role between neural substrates and reduced social function. However, it can be concluded that the dorsal regions found to be related to symptomatology (right hemisphere angular gyrus, supramarginal gyrus, somatosensory association cortex, somatosensory cortex, and dorsolateral prefrontal cortex), are involved in the underlying neural conditions relevant to ASD. Given a well-recognized need for biomarkers for ASD that associate with the clinical phenotype at the individual level, the strong relationships observed between neural activity and both clinician and self-reported social function suggest potential utility in key contexts of use, such as stratification for enrichment of clinical trials (111).

In conclusion, these findings highlight the right dorsal stream system and interactive face processing as regions and tasks of interest for understanding the underlying neural mechanisms that distinguish ASD and TD participants. The specificity of these findings opens new directions for investigating these brain-to-behavior linkages. For example, these regions have previously been implicated in motion sensitivity (48) and raise the interesting hypothesis that reduced face processing in social interactions in ASD is related to reduced sensitivity to the subtle expressive movements of a real face. However, the strong (r = -0.77) negative correlation between social performance (ADOS-2) and the dorsolateral prefrontal cortex in ASD does not fall under that hypothesis and was not predicted. These findings, however, are consistent with face processing activity observed in right lateral prefrontal cortex using fMRI and TD participants (112). Interestingly, in that study it was found that face processing activity observed in the right inferior frontal junction (including regions labeled as DLPFC and frontal eye fields, see Table 3) was primarily responsive to the eyes and not the whole face. Consistent with these prior findings, in the study reported here the strongest negative correlation between ADOS-2 scores and neural activity (-0.77) was in this area and observed during eye contact (Figs 7 and 8). These findings suggest another target of further investigation in this dorsal neural pathway and its role in social performance. Within this framework, our findings are consistent with the hypothesis that eye movements (increased positional variation) during live eye contact and social processing are altered in ASD and suggest that “bottom-up” factors may also impact live eye-to-eye interactions. Altered incoming information due to visual sensing factors such as increased fixation variability could fail to capture higher-order motion cues associated with face and social interactions. Individuals with ASD show raised thresholds for the perception of coherent motion (10, 15, 113). These findings also add support for the “dorsal stream vulnerability” hypothesis in ASD suggesting that mechanisms supporting motion sensitivity such as live face interactions are compromised (48).

### Limitations

The advantages of fNIRS are balanced by technical limitations relative to fMRI. The spatial resolution of fNIRS (approximately 3 cm) does not allow for discrimination of small anatomical differences in functional activity between gyri, and the origin of acquired signals does not extend below the superficial grey matter of the cortex of about 1.5-2.0 cm. Thus, findings of this, and other investigations based on fNIRS technology, are restricted to superficial cortical networks. Although the eye-to-eye contact occurs in a live context which is a novel advance, the gaze situation in the study is relatively constrained which is necessary for experimental control and thus also sets constraints on the investigation of naturally occurring behaviors.

## Data Availability

The datasets analyzed for this study will be made available upon request at fmri.org/ ENDAR and the NIH Data Archive.

http://www.fmri.org

## Author Information and Contributions

Full Name: Joy Hirsch, PhD

Postal Address: Yale School of Medicine, Brain Function Laboratory, 300 George St, Suite 902, New Haven, CT, 06511

Telephone Number: 917.494.7768 Email Address

Contribution(s): Research design and supervision; data analysis; wrote the paper

Full Name: James C. McPartland, PhD

Postal Address: Yale Child Study Center, Nieson Irving Harris Building, 230 South Frontage Road, Floor G, Suite 100A, New Haven, CT, 06519

Telephone Number: 203.785.7179

Contribution(s): Research design and clinical supervision; edited the paper

Full Name: Xian Zhang, PhD

Postal Address: Yale School of Medicine, Department of Psychiatry, Brain Function Laboratory, 300 George St, Suite 902, New Haven, CT, 06511

Telephone Number: 917.494.7768 Email Address

Contribution(s): Analysis of neuroimaging and eye tracking data; edited the paper

Full Name: J. Adam Noah, PhD

Postal Address: Yale School of Medicine, Department of Psychiatry, Brain Function Laboratory, 300 George St, Suite 902, New Haven, CT, 06511

Telephone Number: 917.494.7768 Email Address

Contribution(s): Performed neuroimaging and eye tracking research; analyzed neuroimaging data; maintenance of system calibrations and performance; edited the paper

Full Name: Swethasri Dravida, MD-PhD

Postal Address: Yale School of Medicine, Brain Function Laboratory, 300 George St, Suite 902, New Haven, CT, 06511

Telephone Number: 917.494.7768

Contribution(s): Data acquisition and analysis

Full Name: Adam Naples, PhD

Postal Address: Yale Child Study Center, Nieson Irving Harris Building, 230 South Frontage Road, Floor G, Suite 100A, New Haven, CT, 06519

Telephone Number: 203.785.7179 Email Address

Contribution(s): Research design and clinical supervision

Full Name: Mark Tiede, PhD

Postal Address: Haskins Laboratories, 300 George St, 9^th^ Floor, New Haven, CT, 06511 Telephone Number: 203.865.6163

Contribution(s): Analysis of eye tracking data and manuscript editing

Full Name: Julie M. Wolf, PhD

Postal Address: Yale Child Study Center, Nieson Irving Harris Building, 230 South Frontage Road, New Haven, CT, 06519

Telephone Number: 203.785.5337 Email Address

Contribution(s): Acquired and analyzed clinical data

## Acknowledgements

The content is solely the responsibility of the authors and does not necessarily represent the official views of the National Institutes of Health. All data reported in this paper are available upon request from the corresponding and first author. The authors are grateful to the participants for their essential efforts to advance understanding of ASD; to our two lab partners, CD and IS, for consistent partnership with our participants and the investigators; and to Jen Cuzzocreo for database management and graphical representations of the data.

## Disclosure of Biomedical Financial Interests and Potential Conflicts of Interest

The authors declare that this research was conducted in the absence of any commercial or financial relationships that could be construed as a potential conflict of interest.

## Data Availability Statement

The datasets analyzed for this study will be made available upon request at fmri.org/ENDAR and the NIH Data Archive.

## Supporting Information

**Fig S1.**
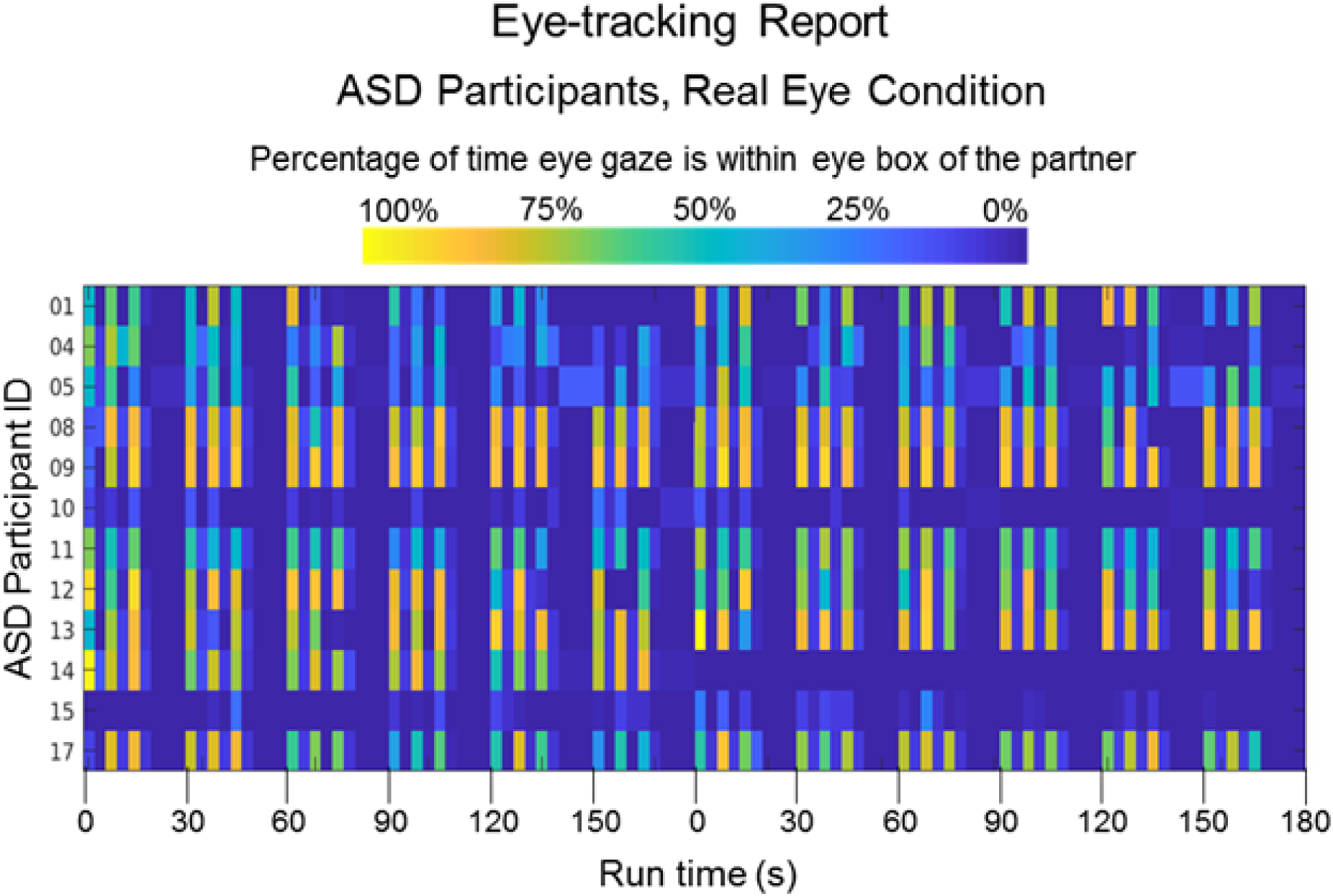
Eye tracking Report for Autism Spectrum Disorder (ASD) Participants, Real Eye Condition. Colors indicate the percentage of time eye gaze is within the eye region of the partner (dark blue = 0% and bright yellow = 100%) during each epoch of the time series (x-axis). The vertical axis includes all ASD participants for whom eye tracking data were acquired.

**Fig S2.**
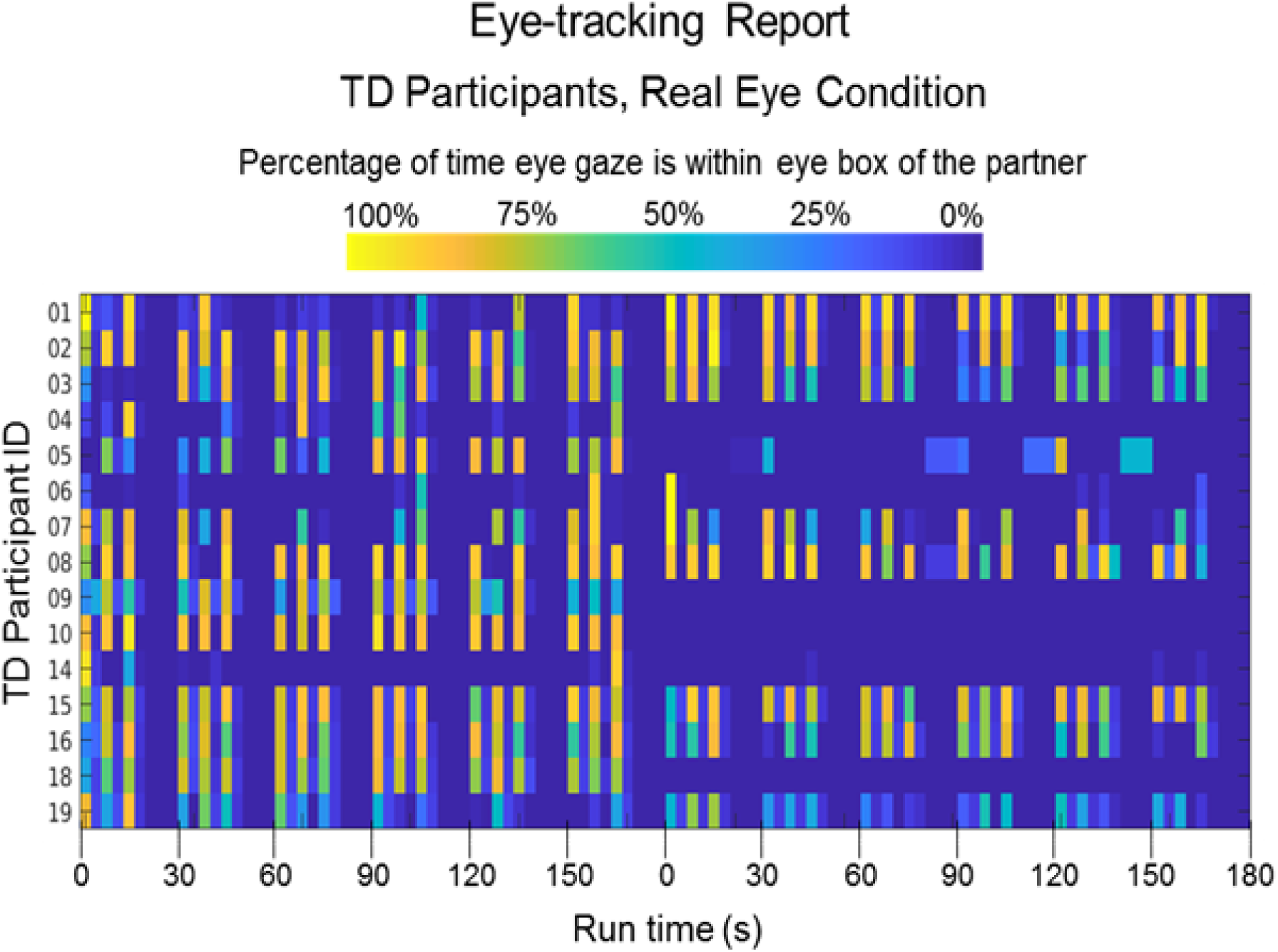
Eye tracking Report for Typically-Developed (TD) Participants, Real Eye Condition. Colors indicate the percentage of time eye gaze is within the eye region of the partner (dark blue = 0% and bright yellow = 100%) during each epoch of the time series (x-axis). The vertical axis includes all TD participants for whom eye tracking data were acquired.

**Table S1.** Demographic information for Autism Spectrum Disorder (ASD) participants. Assessment measures include the Autism-Spectrum Quotient test (AQ, total scores); Broad Autism Phenotype Questionnaire (BAPQ, total scores); Social Responsiveness Scale, Second Edition (SRS-2, raw scores); Beck Anxiety Inventory (BAI, total scores); State-Trait Anxiety Inventory (STAI; total state anxiety scores); Liebowitz Social Anxiety Scale (LSAS, total scores); and the Autism Diagnostic Observation Schedule (ADOS-2, total scores). The Wechsler Abbreviated Scale of Intelligence, 2^nd^ Edition (WASI-II) was administered to estimate full-scale intelligence quotient scores based on four subtests (FSIQ-4). *Indicates data are unavailable.

Study Participants and Behavioral Test Scores: ASD
| ID | M/F | Age | FSIQ-4 | AQ | BAPQ | SRS-2 | BAI | STAI | LSAS | ADOS-2 | Eye tracking |
| --- | --- | --- | --- | --- | --- | --- | --- | --- | --- | --- | --- |
| 1 | Male | 28-32 | 114 | 26 | 123 | 54 | 16 | 21 | 30 | 12 | Yes |
| 2 | Male | 33-37 | 104 | 23 | 121 | 30 | 8 | 31 | 0 | 15 | * |
| 3 | Male | 28-32 | 112 | 30 | 137 | 82 | 11 | 27 | 46 | 16 | * |
| 4 | Male | 23-27 | 108 | 15 | 102 | 53 | 21 | 30 | 56 | 14 | Yes |
| 5 | Male | 23-27 | 97 | 15 | 99 | 41 | 19 | 29 | 43 | 10 | Yes |
| 6 | Male | 23-27 | 90 | 26 | 131 | 132 | 41 | 53 | 79 | 16 | * |
| 7 | Male | 28-32 | 113 | 15 | 94 | 68 | 7 | 44 | 44 | 13 | * |
| 8 | Male | 18-22 | 95 | 24 | 117 | 79 | 24 | 37 | 38 | 15 | Yes |
| 9 | Male | 18-22 | 101 | 30 | 140 | 96 | 27 | 46 | 85 | 18 | Yes |
| 10 | Female | 28-32 | 110 | 20 | 124 | 70 | 9 | 61 | 34 | 17 | Yes |
| 11 | Female | 18-22 | 128 | 32 | 121 | 68 | 4 | 30 | 32 | 9 | Yes |
| 12 | Male | 18-22 | 124 | 25 | 128 | 60 | 1 | 35 | 44 | 9 | Yes |
| 13 | Male | 18-22 | 107 | 14 | 95 | 34 | 9 | 55 | 5 | 12 | Yes |
| 14 | Male | 18-22 | 121 | 30 | 155 | 103 | 17 | 41 | 77 | 11 | Yes |
| 15 | Male | 23-27 | 112 | 20 | 117 | 36 | 3 | 23 | 3 | 13 | Yes |
| 16 | Male | 28-32 | 101 | 47 | 189 | 153 | 25 | 67 | 126 | 12 | * |
| 17 | Female | 28-32 | 109 | 23 | 94 | 65 | 15 | 33 | 38 | 11 | Yes |

\*Data unavailable

**Table S2.** Demographic information for Typically-Developed (TD) participants. Assessment measures include the Autism-Spectrum Quotient test (AQ, total scores); Broad Autism Phenotype Questionnaire (BAPQ, total scores); Social Responsiveness Scale, Second Edition (SRS-2, raw scores); Beck Anxiety Inventory (BAI, total scores); State-Trait Anxiety Inventory (STAI; total state anxiety scores); and the Liebowitz Social Anxiety Scale (LSAS, total scores). The Wechsler Abbreviated Scale of Intelligence, 2^nd^ Edition (WASI-II) was administered to estimate full-scale intelligence quotient scores based on four subtests (FSIQ-4). *Indicates data are unavailable.

Study Participants and Behavioral Test Scores: TD
| ID | M/F | Age | FSIQ-4 | AQ | BAPQ | SRS-2 | BAI | STAI | LSAS | Eye tracking |
| --- | --- | --- | --- | --- | --- | --- | --- | --- | --- | --- |
| 1 | Male | 18-22 | * | 12 | 77 | 52 | 10 | * | 23 | Yes |
| 2 | Male | 18-22 | 123 | 23 | 118 | 55 | 4 | 39 | 52 | Yes |
| 3 | Male | 28-32 | 113 | 6 | 85 | 9 | 4 | 26 | 24 | Yes |
| 4 | Male | 23-27 | * | * | 72 | 16 | * | * | * | Yes |
| 5 | Female | 23-27 | 129 | 6 | 70 | 20 | 1 | 25 | 23 | Yes |
| 6 | Female | 23-27 | 114 | 8 | 79 | 27 | 5 | 28 | 43 | Yes |
| 7 | Female | 28-32 | 103 | 1 | 53 | 6 | 3 | 21 | 22 | Yes |
| 8 | Male | 28-32 | * | 26 | 109 | 48 | 0 | 23 | 23 | Yes |
| 9 | Female | 18-22 | 79 | 32 | 121 | 64 | 5 | 37 | 44 | Yes |
| 10 | Female | 23-27 | 119 | 22 | 111 | 37 | 26 | 62 | 60 | Yes |
| 11 | Female | 28-32 | 107 | 14 | 88 | 36 | 2 | 29 | 63 | * |
| 12 | Male | 28-32 | 111 | 10 | 73 | 25 | 5 | 28 | 27 | * |
| 13 | Female | 23-27 | 126 | 14 | 93 | 43 | 10 | 38 | 12 | * |
| 14 | Male | 18-22 | * | 25 | 105 | 84 | 7 | 27 | 59 | Yes |
| 15 | Female | 23-27 | 122 | 7 | 79 | 12 | 0 | 26 | 32 | Yes |
| 16 | Male | 38-42 | * | 21 | 124 | 63 | 9 | 56 | 56 | Yes |
| 17 | Male | 18-22 | * | 19 | 96 | 45 | 0 | 22 | 45 | * |
| 18 | Male | 28-32 | 119 | 19 | 89 | 38 | 12 | 26 | 46 | Yes |
| 19 | Male | 38-42 | 121 | 10 | 73 | 31 | 0 | 20 | 18 | Yes |

\*Data unavailable

**Table S3.** Comparison of Autism Spectrum Disorder (ASD) and Typically-Developed (TD) participant groups by gender, handedness, and age. Groups were similar in terms of age and handedness; however, the ratio of male to female participants was higher in the ASD group than in the TD group. The gender composition of the ASD group is consistent with the estimated 4:1 male:female ratio of ASD diagnosis. This ratio increases to 6 males diagnosed with ASD for every 1 female in people whose cognitive functioning is within or above normal limits, such as those in our sample (Kirkovski, M., Enticott, P. G., & Fitzgerald, P. B. (2013). A review of the role of female gender in autism spectrum disorders. Journal of Autism and Developmental Disorders, 43(11), 2584-2603).

Comparison of ASD and TD Groups by Gender, Handedness, and Age
|  | ASD | TD | n | % |
| --- | --- | --- | --- | --- |
| **Female** | 3 | 8 | 11 | 31% |
| **Male** | 14 | 11 | 25 | 69% |
| **n** | 17 | 19 | **36** |  |
| **%** | 47% | 53% |  |  |
| **Handedness** |  |  |  |  |
| Right | 12 | 18 | 30 | 83.3% |
| Left | 3 | 0 | 3 | 8.3% |
| Ambidextrous | 2 | 1 | 3 | 8.3% |
| **Age (years)** |  |  |  |  |
| Mean | 25 | 26 |  |  |
| Median | 26 | 26 |  |  |
| Range | 18-34 | 19-38 |  |  |
| SD | ± 4.9 | ± 5.8 |  |  |

**Table S4.** Statistical comparisons (independent t-tests, two-tailed assuming unequal variances) of scores between Typically-Developed (TD) and Autism Spectrum Disorder (ASD) groups are consistent with differences for the Autism-Spectrum Quotient test; Broad Autism Phenotype Questionnaire; Social Responsiveness Scale, 2^nd^ Edition; and the Beck Anxiety Inventory. No evidence was found for differences between the groups for FSIQ-4 (estimated by the Wechsler Abbreviated Scale of Intelligence); State-Trait Anxiety Inventory (state anxiety items only); or the Liebowitz Social Anxiety Scale, and is taken as evidence in favor of matched groups with respect to these metrics.

**Statistical Comparisons of Clinical and Behavioral Assessments for ASD and TD Groups**
| Measure | p-value |
| --- | --- |
| Autism-Spectrum Quotient | 0.002 |
| Broad Autism Phenotype Questionnaire | 0.002 |
| Social Responsiveness Scale | 0.002 |
| Beck Anxiety Inventory | 0.016 |
| State-Trait Anxiety Inventory | 0.067 |
| Liebowitz Social Anxiety Scale | 0.363 |
| Wechsler Abbreviated Scale of Intelligence (FSIQ-4) | 0.539 |

**Table S5.**
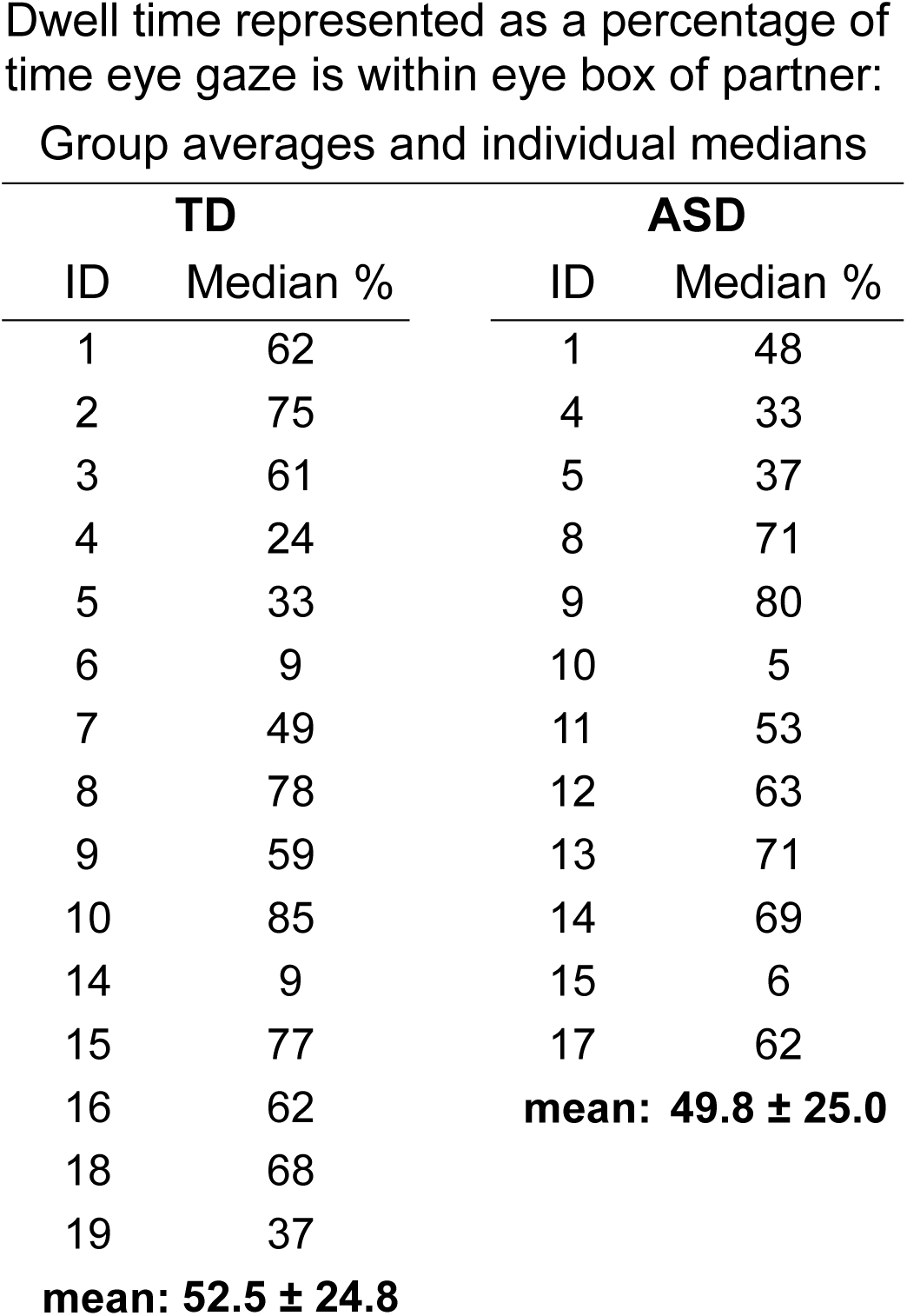
Group averages and individual median percentages of eye-gaze time within the eye box of partners for typically developed (TD) participants (left column) and autism spectrum disorder (ASD) participants (right column) during the Real Eye Condition. A t-test of these median percentages shows t(25) = 0.28 n.s. See S1 and S2 Figs for a graphical run-by-run representation of eye tracking performance.

